# Plasma Proteogenomics Identifies RGMB as a Candidate for Primary Open-Angle Glaucoma Detected Only in East Asians

**DOI:** 10.64898/2026.10.26.26366459

**Authors:** Shuzo Okuno, Shiro Mizoue, Koji Namiguchi, Masashi Yukawa, Atsushi Shiraishi

## Abstract

**Purpose:** In East Asians most primary open-angle glaucoma (POAG) occurs at statistically normal intraocular pressure (IOP). We sought plasma proteins linked to POAG through genetic signal not shared with IOP.

**Methods:** We integrated the BioBank Japan POAG GWAS with a Japanese plasma pQTL resource (Nagahama; SomaScan v4) using SMR/HEIDI and colocalization against POAG and IOP datasets, including a Genomic-SEM–derived non-IOP POAG component, adding a symmetric proteome-wide cross-ancestry colocalization, fine-mapping and multi-signal colocalization of the RGMB region, a formal heterogeneity test, and expression mapping in retina and trabecular meshwork.

**Results:** Of 679 plasma proteins, four colocalized with POAG in East Asians only, five in Europeans only, none in both. SMR nominated RGMB as a suggestive risk factor (b_SMR = +0.277; HEIDI P > 0.05; PP4 = 0.684; rs1037599); none survived Bonferroni correction. RGMB did not colocalize with European IOP, a European non-IOP POAG component, or European POAG on the same platform, despite a common, directionally consistent pQTL instrument. Multi-signal colocalization reproduced both the East Asian signal and the European null. The per-allele POAG effect differed between ancestries (Cochran’s Q P = 0.0036; I² = 88%), the better-powered European data excluding an effect of East Asian magnitude. RGMB and its receptors were expressed in trabecular meshwork cells and unchanged by TGF-β2.

**Conclusions:** This triangulation nominates the RGMB–neogenin–BMP axis as a POAG candidate detected in East Asians but not Europeans, which does not colocalize with IOP-related traits. The findings are exploratory; ancestry-matched replication and functional validation are required.

## Introduction

Primary open-angle glaucoma (POAG) is a leading cause of irreversible blindness, characterized by progressive loss of retinal ganglion cells (RGCs) and corresponding visual-field deficits. Elevated intraocular pressure (IOP) is the principal modifiable risk factor, and current standard of care centers almost entirely on lowering IOP pharmacologically or surgically. However, a substantial fraction of patients continue to progress despite well-controlled IOP, and the relative contribution of pressure-independent mechanisms differs markedly across populations.

This heterogeneity is especially evident in East Asia. In European-ancestry populations, POAG is more often accompanied by ocular hypertension: across population-based studies the proportion of NTG among open-angle glaucoma in white patients is comparatively low (approximately 30–39%^1^), so elevated-IOP (“high-tension”) POAG predominates^2^ (the Italian Egna-Neumarkt Study reported an NTG proportion of about 30%^3^). By contrast, population-based screening in Japan has shown that the majority of POAG cases occur at statistically normal IOP. The Tajimi Study^4^ reported that approximately 92% of Japanese POAG patients had IOP within the normal range (≤ 21 mmHg) at detection, consistent with a normal-tension glaucoma (NTG) phenotype, and NTG comprises the majority (roughly 52–92%) of open-angle glaucoma across Asian epidemiological studies^2^; a high prevalence of NTG was also observed in the Hisayama Study^5^. This contrast — high-tension POAG predominating in Europeans versus NTG predominating in the Japanese — indicates that pressure-independent processes contribute substantially to optic neuropathy in East Asian populations, and motivates an ancestry-aware search for IOP-independent molecular drivers, which remain incompletely defined.

Although GWAS have identified more than one hundred POAG susceptibility loci, many implicated variants are shared with IOP, and large discovery cohorts have been dominated by individuals of European ancestry, in whom high-tension glaucoma is comparatively more common. Analytical approaches such as Genomic SEM can partition POAG genetic signal into IOP-dependent and IOP-independent (“non-IOP”) components^6^, but such models remain bounded by the ancestry of their input GWAS. Proteogenomic integration offers a complementary, data-driven route from genetic association toward candidate causal biology by using pQTLs as instruments for circulating protein intermediaries. Several plasma proteome-wide MR and colocalization analyses of POAG have already been reported using European pQTL resources (deCODE and UKB-PPP) against European POAG GWAS ^7,8^ nominating proteins such as SVEP1, ITIH1 and TMEM190; however, these analyses were confined to European populations, and Jia et al. explicitly noted the need to test whether such findings extend to East Asian or African ancestries. A symmetric, like-for-like comparison of the plasma proteogenomic profile of POAG between East Asian and European populations has not, to our knowledge, been reported.

Here we integrated the BBJ POAG GWAS with a Japanese plasma pQTL resource (Nagahama Study) using SMR, and triangulated prioritized candidates through multi-dataset colocalization, comparison with a European non-IOP POAG component, and single-cell retinal expression mapping. We prioritize plasma RGMB as a candidate IOP-independent risk factor for POAG. We emphasize at the outset that no protein survived multiple-testing correction; the results are therefore exploratory and hypothesis-generating, and are intended to nominate a pathway for ancestry-matched replication and functional study rather than to establish causality. We note at the outset that the East Asian–enriched interpretation rests on a single East Asian discovery cohort (BioBank Japan POAG); no independent East Asian POAG outcome dataset is currently available in the public domain for replication, so this remains a candidate interpretation pending ancestry-matched confirmation.

## Methods

### Data sources

This study was a secondary analysis of publicly available, summary-level data; no individual-level data were accessed and no new human or animal experiments were performed. Institutional review board or equivalent ethical approval and informed consent from participants were obtained in each of the original studies, as described in their respective primary publications. The study adhered to the tenets of the Declaration of Helsinki.

The exposure datasets were the BBJ POAG GWAS^9^ (3,980 cases and 18,815 controls; originally in genome build hg19) and a Japanese plasma *cis*-pQTL resource from the Nagahama Study^10^ (SomaScan v4 aptamer panel; GRCh38). BBJ summary statistics were lifted from hg19 to GRCh38 with pyliftover prior to analysis. Linkage-disequilibrium (LD) estimation used a 1000 Genomes East Asian (EAS) reference panel^11^ (*n* = 585).

### Definition of cis-pQTL instruments

A *cis*-pQTL was defined as a variant within ±1 Mb of the gene encoding the assayed protein that was associated with plasma levels of that protein at genome-wide significance (*P* < 5×10⁻⁸). For each protein, association was assessed by regressing measured plasma protein level on variant genotype with study covariates; the test statistic is the effect estimate divided by its standard error, and because the standard error scales approximately with the inverse square root of the sample size, a given true effect is more likely to reach significance in a larger sample. The Nagahama resource was generated on the SomaScan v4 platform (4,392 aptamers analyzed, corresponding to 4,239 unique proteins), whereas the European deCODE resource assays a partially overlapping SomaScan v4 panel (4,907 aptamers); the number of proteins with at least one genome-wide-significant *cis*-pQTL was therefore far larger in the much larger deCODE sample (*n* ≈ 35,559) than in the Nagahama sample (*n* ≈ 1,823), which yielded usable *cis* instruments for 679 aptamers. This difference reflects statistical power to detect *cis*-pQTLs, not a difference in the proteins measured. Instruments were restricted to the *cis* window to limit horizontal pleiotropy, and where multiple variants were available the analyses used the lead (most significant) *cis* variant for SMR and the full set of overlapping *cis* variants for colocalization. In the source resource^10^ a variant was classified as *cis* when located within the protein-coding gene extended by 500 kb upstream of the gene start and 500 kb downstream of the gene end; conditionally independent signals were obtained by GCTA-COJO (--cojo-p 5×10⁻⁸) and the study-wide significance threshold was *P* < 1.14×10⁻¹¹ (5×10⁻⁸ divided by 4,392 assayed aptamers). The publicly released resource comprised 679 lead *cis*-pQTLs, which constituted the exposure panel for the present analysis.

### Summary-data-based Mendelian randomization

SMR (v1.3.1)^12^ with the HEIDI test was used to screen plasma proteins for evidence of a shared genetic signal with POAG, using *cis*-pQTL instruments. A non-significant HEIDI result (*P* > 0.05) was interpreted as a lack of evidence that the SMR association was driven by linkage with a distinct nearby variant. Because no protein passed Bonferroni correction for the number of proteins tested, prioritization was treated as suggestive and hypothesis-generating throughout.

### Bayesian colocalization

Colocalization was performed with coloc (coloc.abf, default priors)^13^. pQTL and quantitative outcome datasets were analyzed under the quantitative-trait model. To integrate the EAS pQTL data (GRCh38) with outcome datasets reported in differing builds, variants were matched by rsID, which is build-independent; coloc.abf is invariant to allele orientation under the default implementation. For each test we report the posterior probabilities for distinct causal variants (PP.H3) and a shared causal variant (PP.H4; hereafter PP4), and note the dominant hypothesis (e.g., PP.H2, a signal in only one dataset).

Colocalization inputs and prior sensitivity. For every colocalization the trait type was specified explicitly. Plasma pQTL datasets were analyzed as quantitative traits (type = “quant”) with the sample size of the corresponding pQTL resource (Nagahama n = 1,823; deCODE n = 35,559; China Kadoorie Biobank n ≈ 3,974). POAG datasets were analyzed as case–control traits (type = “cc”) with the total sample size and the case fraction supplied: Biobank Japan, N = 22,795 with s = 0.175 (3,980 cases and 18,815 controls); the European meta-analysis of Gharahkhani et al., N = 216,257 with s = 0.0771 (16,677 cases and 199,580 controls). Quantitative outcome datasets (IOP; the non-IOP latent component) were analyzed under the quantitative model with the sample sizes given above. Approximate Bayes factors were computed from the per-variant effect estimate and its standard error; minor allele frequencies, where required, were taken from the ancestry-matched pQTL resource, and the variance of the quantitative exposure was set to unity because the plasma protein levels had been rank-normalized in the source resources. Every colocalization reported here was repeated at prior probabilities of shared causality p12 = 1 × 10⁻⁵ (the coloc default), 1 × 10⁻⁶ and 1 × 10⁻⁷; posteriors at all three priors are given in Supplementary Table S4. The number of variants contributing to each test is reported alongside every posterior.

Multi-signal colocalization. Because coloc.abf assumes at most one causal variant per trait within a region, and because fine-mapping resolved multiple independent signals in the European pQTL data, we repeated the RGMB colocalization with methods that accommodate multiple signals. Ancestry-matched LD was taken from the same 1000 Genomes Phase 3 panels used for fine-mapping, with correlations signed to the effect allele of each summary-statistic dataset. Variants whose z-scores were inconsistent with the reference LD were identified with the kriging_rss diagnostic of susieR (|z| > 2 together with logLR > 2) and removed over two iterations; this reduced the summary-statistic–LD consistency estimate (estimate_s_rss) from 0.80 to 0.10 in the East Asian arm and from 0.62 to 0.09 in the European arm. SuSiE fine-mapping was then repeated on the cleaned variant sets, and colocalization was evaluated with coloc.signals in both conditioning and masking modes. All analyses were run across cis-windows of ±150, ±250 and ±500 kb around the lead variant and with ridge regularization of the LD matrix (λ = 0 for the primary analysis and λ = 0.01 as a sensitivity analysis), each at the three priors above. Because SuSiE applied to the POAG side returned no 95% credible set in either ancestry, reflecting the limited power of the outcome GWAS at this locus, coloc.susie could not be used, and conditioning and masking were used in its place.

Formal test of cross-ancestry heterogeneity. To test ancestry heterogeneity directly rather than by thresholding the colocalization posterior, we compared the per-allele POAG effect of rs1037599 between Biobank Japan and the European meta-analysis using Cochran’s Q with one degree of freedom, together with I² and an equivalent Z test of the difference between the two estimates. The same test was applied to the 22 East Asian LD proxies of the lead variant (r² ≥ 0.8 in the 1000 Genomes East Asian panel); all effects were aligned to the plasma-RGMB-raising allele, and P values across proxies were adjusted by the Benjamini–Hochberg procedure. Because the two proteomic platforms report on different scales, no heterogeneity test was applied to the exposure (pQTL) estimates; instrument strength was instead compared as z statistics. To quantify the asymmetry in power we report, for the European dataset, the standard error, the 95% confidence interval, the smallest effect detectable at α = 0.05, and the test statistic that the East Asian point estimate would have produced had it been present in the European data.

### Cross-ancestry proteome-wide colocalization scan

To place the RGMB signal in a systematic context, we performed a symmetric, proteome-wide colocalization comparison between ancestries on an identical platform (SomaScan v4) and with an identical method (coloc.abf, default priors). For each of the 679 plasma proteins with a significant *cis*-pQTL instrument in the Nagahama resource (the common panel defining the comparison: the 679 lead *cis*-pQTLs released by the Nagahama resource; both resources use the SomaScan v4 platform — Nagahama analyzing 4,392 aptamers and deCODE 4,907 — and the 679 reflects the instruments reaching significance in the smaller Nagahama sample), we tested colocalization with POAG in East Asians (Nagahama *cis*-pQTL × BBJ POAG) and, in parallel, in Europeans (deCODE *cis*-pQTL^14^, *n* = 35,559 × European POAG^15^). Variants were matched on GRCh38 position; outcome effects were aligned to the deCODE effect allele; minor allele frequencies were taken from the pQTL data; and deCODE multiallelic mis-encodings (effect allele = other allele) were removed. Of the 679 East Asian aptamers, 678 had a matching deCODE file. Each protein was classified by its posterior probability of a shared causal variant (PP4) as East Asian–enriched (PP4 ≥ 0.5 in East Asians and < 0.5 in Europeans), European–enriched (the reverse), shared (≥ 0.5 in both), or neither.

### IOP and non-IOP POAG datasets

IOP comparators were the IGGC IOP GWAS^16^ (GWAS Catalog accession GCST009413; *n* = 31,269; European ancestry) and the Pan-UKBB European IOP GWAS (corneal-compensated IOP, UK Biobank field 5254; *n* = 87,199; rsIDs attached via the Pan-UKBB variant manifest^17^). The IOP-independent POAG component was the non-IOP latent factor generated by Genomic SEM GWAS-by-subtraction ^6^ (deposited on Zenodo), in which European IOP genetic effects were subtracted from European POAG; this latent factor was analyzed under the quantitative model with a nominal effective sample size. Lead loci from the Japanese Tohoku Medical Megabank (TMM) IOP GWAS^18^ (*n* = 33,452) were used for a look-up of the RGMB region. For a within-ancestry test, we additionally colocalized a European RGMB *cis*-pQTL from deCODE (SomaScan v4; the same aptamer, SeqId 3331-8, as the East Asian resource; *n* = 35,559) with the European POAG meta-analysis of Gharahkhani et al.^15^ (GWAS Catalog accession GCST90011766; 16,677 cases and 199,580 controls), restricting both datasets to the RGMB *cis*-window (lead variant ± 1 Mb) before rsID-based matching. The population frequency of the lead variant rs1037599 was compared between East Asian (Nagahama pQTL) and European (Pan-UKBB) data.

### Cross-ancestry fine-mapping and European POAG look-up

To probe the basis of the East Asian–detected signal, we fine-mapped the RGMB *cis*-region (chr5:97,768,004–99,798,643, GRCh38; ±1 Mb of the gene body) separately in the East Asian (Nagahama) and European (deCODE) plasma pQTL datasets using the sum-of-single-effects model^19^ (SuSiE; susieR, susie_rss, maximum number of signals *L* = 10, residual variance fixed). Ancestry-matched LD was estimated from 1000 Genomes Phase 3 reference panels (East Asian, *n* = 504; European, *n* = 503), restricted to the 998 variants shared between the two pQTL datasets. Concordance between each reference panel and the corresponding summary statistics was assessed with the consistency diagnostic of susieR (East Asian λ = 0.088; European λ = 0.072).

For each ancestry we recorded the 95% credible sets and the variant with the highest posterior inclusion probability (PIP), and summarized cross-ancestry LD as the squared correlation (r²) between the lead variant rs1037599 and the pQTL credible-set variants within each panel. We additionally attempted fine-mapping of the BBJ POAG GWAS over the same variant set (effective *n* = 13,140; variants matched to the reference panel by GRCh37 position). Finally, the East Asian credible-set variants, rs1037599, and the European-dominant RGMB regulator were looked up by rsID in the European POAG GWAS (Gharahkhani et al.; GCST90011766) to evaluate their association in Europeans.

### Regulatory annotation and expression look-ups

To explore the regulatory basis of the East Asian signal, the lead variant rs1037599 and its East Asian LD proxies (r² ≥ 0.8 in the 1000 Genomes East Asian panel, *n* = 504) were annotated for regulatory potential using RegulomeDB^20^ (rank) and ENCODE SCREEN^21^ candidate *cis*-regulatory elements (cCRE class), and were screened for allele-specific transcription-factor motif changes with SNP2TFBS^22^; pairwise LD (r²) among these variants and with the lead was computed in the same panel with PLINK (Supplementary Figure S4 and Supplementary Note). As orthogonal expression evidence, rs1037599 and rs1563318 were queried as *cis*-eQTLs for RGMB in GTEx v10, recording the normalized effect size (NES) and *P* value of each significant single-tissue eQTL with the NES oriented to the plasma-RGMB-raising allele. Allele frequencies of the RGMB-raising allele in East Asian and non-Finnish European populations were obtained from gnomAD v4^23^. These annotations are exploratory and hypothesis-generating; no ocular tissue is represented in the expression resources.

### Single-cell retinal expression

Cell-type expression was examined in the CZ CELLxGENE Discover human retina atlas^24^ (∼4.8 million cells). For each gene and retinal cell type we computed the percentage of cells with detected expression (number of expressing cells divided by cell count) and the scaled mean expression. We examined RGMB together with pathway partners NEO1 (neogenin), RGMA, BMP2, and BMPR1A; the RGC marker RBPMS served as a positive control for atlas labeling and scaling.

Trabecular meshwork expression. Because the conventional outflow pathway is central to POAG pathophysiology, we examined RGMB-pathway expression in two independent public RNA-seq datasets of primary human trabecular meshwork (TM) cells: GSE236302 (eight donors, untreated control versus TGF-β2 treatment; seven donors contributed both conditions) and GSE195756 (eight samples). Counts were normalized to counts per million (CPM), and the abundance of each gene was expressed both as CPM and as a percentile rank among the 15,973 genes with mean CPM > 1, so that abundance is judged against the expressed transcriptome rather than against all annotated genes. TM identity was verified with MYOC, CHI3L1, MGP, TIMP3 and DCN, and the response to TGF-β2 was verified with SERPINE1, ACTA2, COL1A1 and FN1. Paired comparisons used paired t tests on log2 CPM across the seven donors contributing both conditions, with Benjamini–Hochberg correction across the gene panel examined. GSE195756 was used only to confirm expression, because its differential-expression contrast showed no response in the canonical mechanoresponsive genes (CCN1, CCN2, ANKRD1) and is therefore uninformative about stimulus response.

### Independent East Asian instrument validation (China Kadoorie Biobank)

To validate the RGMB cis-pQTL instrument in an independent East Asian cohort, we used plasma proteomic GWAS summary statistics from the China Kadoorie Biobank (CKB)^25^, a Chinese cohort assayed with the antibody-based Olink Explore platform^26^, accessed through the public CKB PheWeb portal (https://pheweb.ckbiobank.org). For the RGMB locus we retrieved the regional Olink cis-pQTL statistics (GRCh38) and identified the discovery lead variant rs1037599. Effect-direction concordance was evaluated against the Nagahama SomaScan discovery data and the European deCODE SomaScan statistics by orienting all estimates to the RGMB-increasing allele; because Olink (normalized protein expression) and SomaScan (relative fluorescence units) report on different scales, only the effect allele and the sign of the effect were compared. We then repeated colocalization (coloc.abf) between the CKB RGMB cis-pQTL and the Biobank Japan POAG GWAS. Because the CKB statistics are on GRCh38 whereas the Biobank Japan summary data are on GRCh37/hg19, variants were matched by rsID (build-independent), yielding 976 shared variants in the cis region. Robustness to the prior probability of shared causality was assessed by repeating coloc.abf at p12 = 1 × 10⁻⁵ (default), 1 × 10⁻⁶, and 1 × 10⁻⁷. Because the outcome dataset is identical to the discovery analysis, this procedure constitutes validation of the instrument (an independent East Asian cis-pQTL) rather than replication of an independent outcome GWAS.

### Software

Analyses used SMR v1.3.1, the R package coloc (v5 series), the R package susieR for statistical fine-mapping (susie_rss), PLINK^27^ with 1000 Genomes Phase 3 East Asian and European reference panels for LD, pyliftover for build conversion, and standard R/Python tooling for data handling and visualization.

## Results

### Cross-ancestry comparison of plasma colocalization profiles

Across all 679 plasma proteins, the colocalization profiles of the two ancestries were largely non-overlapping (Figure 1; Tables 1 and 2). Four proteins met the colocalization threshold (PP4 ≥ 0.5) in East Asians only — LILRB2 (PP4 = 0.878), LILRB5 (0.878), TMEM190 (0.738) and RGMB (0.684) — whereas five did so in Europeans only — ITIH1 (0.749), ADPRHL2 (0.655), PLXNA1 (0.546), SVEP1 (0.545) and MMAB (0.501). No protein colocalized with POAG in both ancestries. Among the East Asian–enriched proteins, RGMB was one of the most biologically coherent candidates for an intraocular-pressure–independent mechanism, given the role of the RGM–neogenin–BMP axis in retinal ganglion cell axon guidance; by contrast, LILRB2 and LILRB5 are immune receptors mapping to the LD-complex chromosome-19 leukocyte immunoglobulin-like receptor cluster and were supported by fewer shared variants (686 and 690, versus 1,010 for RGMB), and TMEM190 has no established function. Notably, although LILRB2 and LILRB5 showed higher colocalization posterior probabilities than RGMB, their PP4 values were nearly identical (0.8784 and 0.8784), consistent with the two highly homologous, closely linked genes capturing a shared regional signal rather than two independent shared causal variants; in a high-LD, copy-number-variable locus the absolute PP4 should not be read in isolation. Of the two, only LILRB5 passed the SMR/HEIDI filter, and its HEIDI test rested on only five variants (P = 0.48), giving it little power to detect heterogeneity; LILRB2 was not among the proteins passing that filter (Supplementary Table S9). RGMB, in contrast, was supported concordantly by SMR, a non-significant HEIDI test based on 20 variants, and colocalization, making it the more interpretable candidate despite its lower PP4. We therefore carried RGMB forward as an exemplar for detailed characterization. We note that no protein reached PP4 > 0.8 in Europeans, so the absence of shared signals is observed at moderate posterior probabilities and may reflect ancestry-differential effects, differences in LD, or differences in statistical power rather than a definitive biological dichotomy.

**Figure 1.**
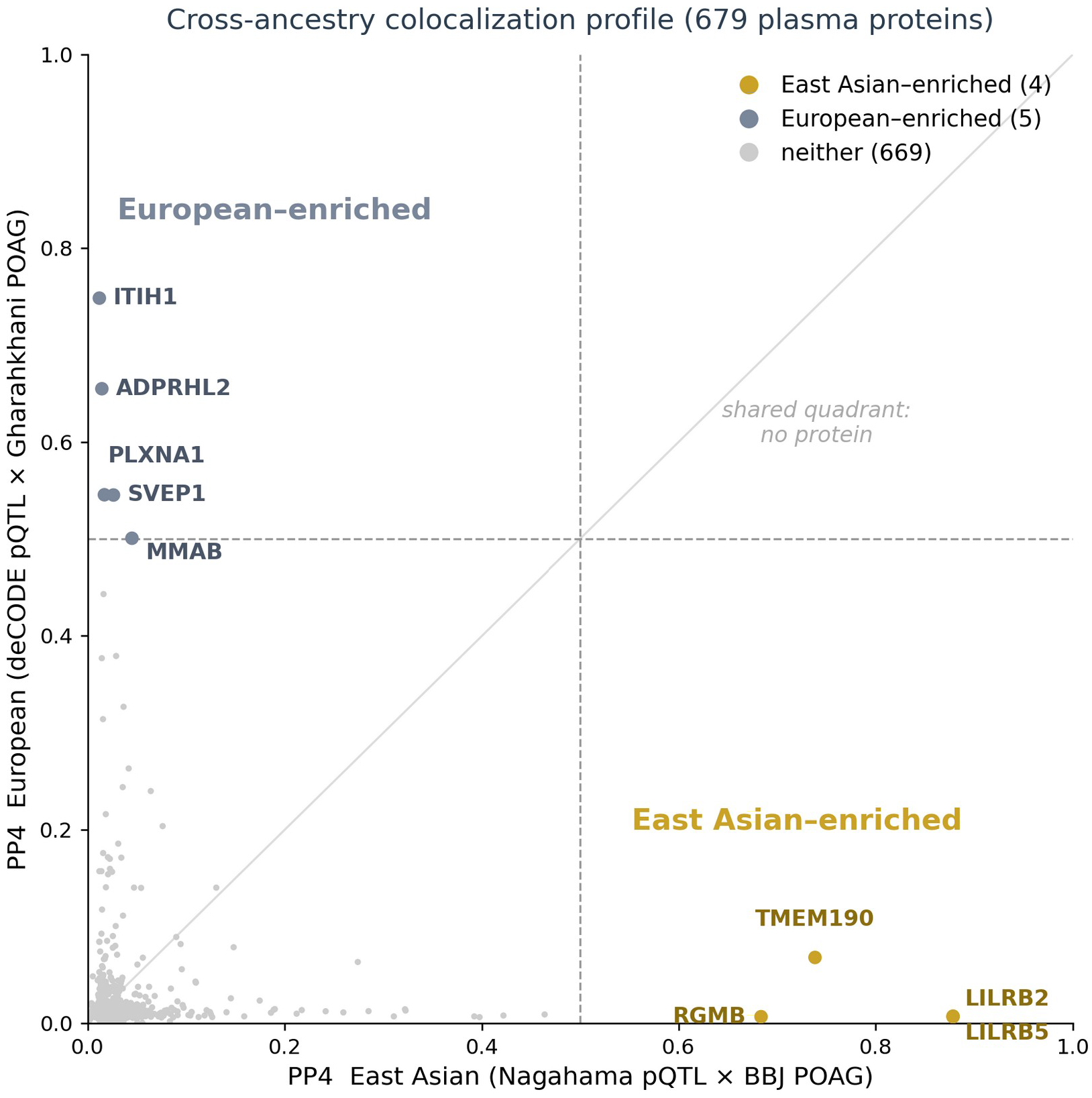
Cross-ancestry colocalization profile of 679 plasma proteins for POAG. Each point is one plasma protein, plotted by its posterior probability of a shared causal variant with POAG (PP4) in East Asians (Nagahama *cis*-pQTL × BBJ POAG; x-axis) versus Europeans (deCODE *cis*-pQTL × Gharahkhani POAG; y-axis), using the same platform (SomaScan v4) and method (coloc.abf). Dashed lines mark PP4 = 0.5. Four proteins are East Asian–enriched (gold; lower right, including RGMB), five are European–enriched (slate; upper left), and none fall in the shared (upper-right) quadrant.

**Table 1.**
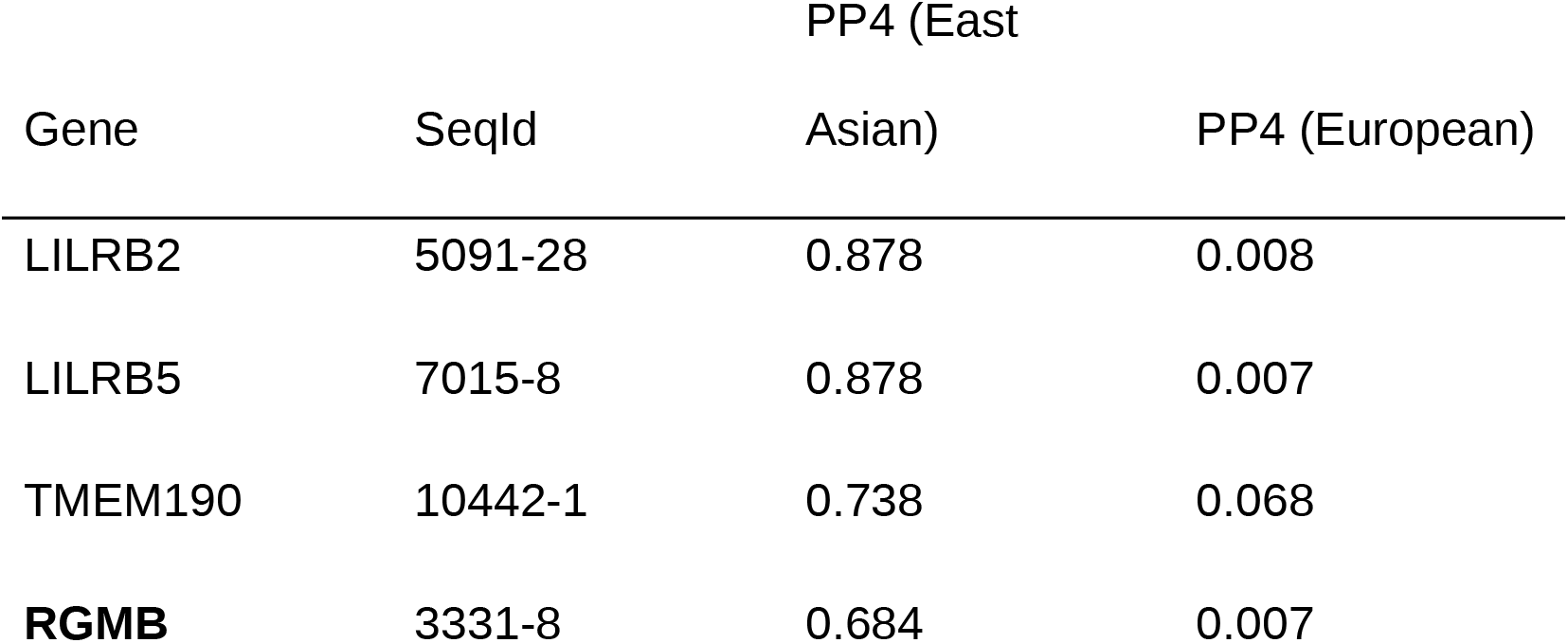
Plasma proteins colocalizing with POAG in East Asians only (PP4 ≥ 0.5 in East Asians, < 0.5 in Europeans).

| Gene | SeqId | PP4 (East Asian) | PP4 (European) |
| --- | --- | --- | --- |
| LILRB2 | 5091-28 | 0.878 | 0.008 |
| LILRB5 | 7015-8 | 0.878 | 0.007 |
| TMEM190 | 10442-1 | 0.738 | 0.068 |
| **RGMB** | 3331-8 | 0.684 | 0.007 |

**Table 2.**
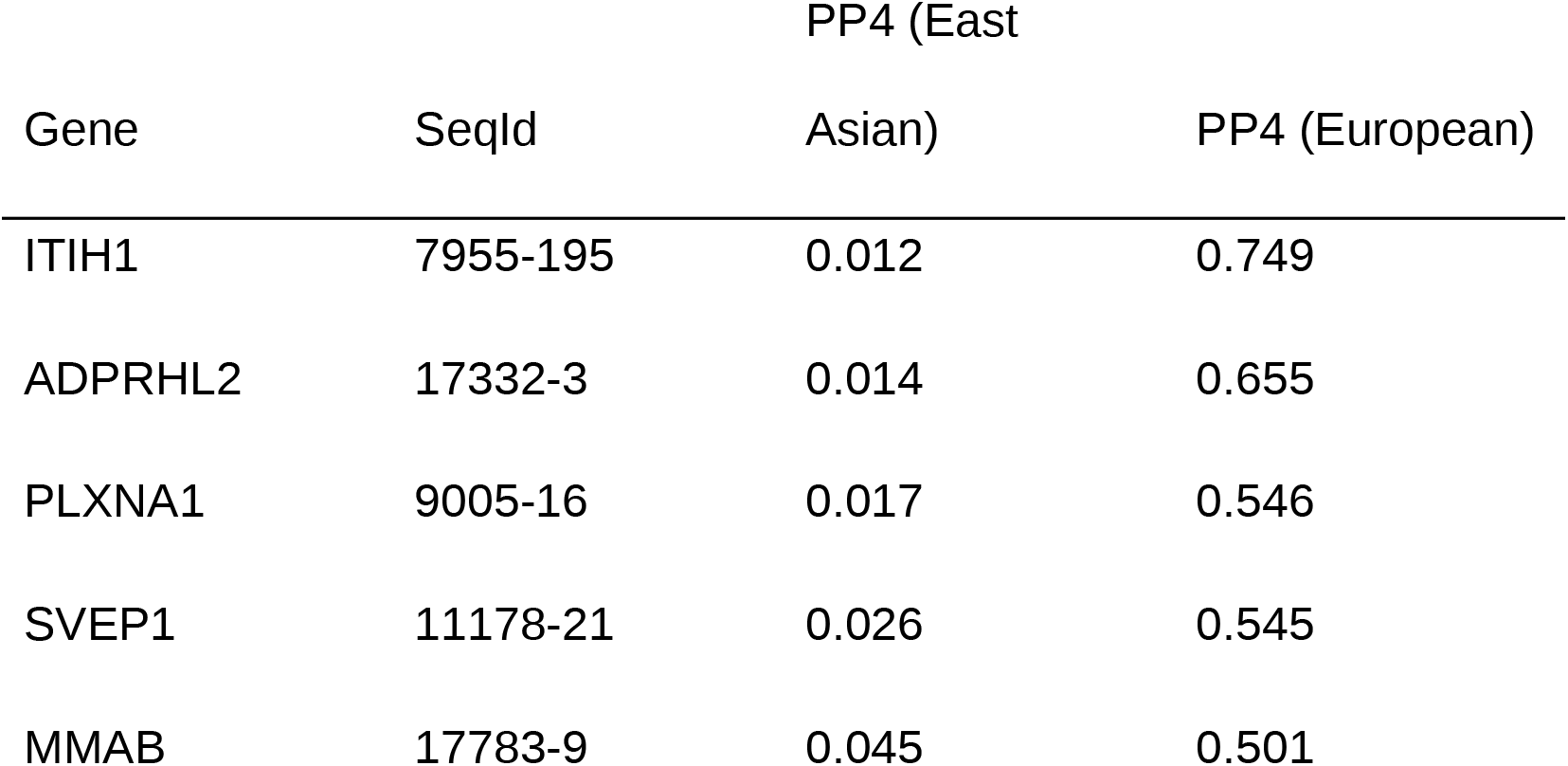
Plasma proteins colocalizing with POAG in Europeans only (PP4 ≥ 0.5 in Europeans, < 0.5 in East Asians).

| Gene | SeqId | PP4 (East Asian) | PP4 (European) |
| --- | --- | --- | --- |
| ITIH1 | 7955-195 | 0.012 | 0.749 |
| ADPRHL2 | 17332-3 | 0.014 | 0.655 |
| PLXNA1 | 9005-16 | 0.017 | 0.546 |
| SVEP1 | 11178-21 | 0.026 | 0.545 |
| MMAB | 17783-9 | 0.045 | 0.501 |

### Proteogenomic prioritization of RGMB

Integrating the BBJ POAG GWAS with the Nagahama plasma *cis*-pQTL resource by SMR, plasma RGMB emerged as a suggestive risk factor for POAG (Figure 2), with a positive estimated effect on susceptibility (*b*_SMR_ = +0.277; SE 0.093; *P* = 0.0029) and a non-significant HEIDI test (*P* = 0.95; 20 variants), the latter providing no evidence that the association was attributable to linkage with a separate confounding variant. Bayesian colocalization provided moderate but not definitive support for a shared signal between plasma RGMB and POAG (PP4 = 0.684; lead variant rs1037599, chr5:98,447,787, GRCh38). We note that PP4 = 0.684 is below the conventional 0.8 threshold for strong colocalization, and that no protein in the screen survived Bonferroni correction; RGMB is therefore presented as a prioritized candidate rather than an established causal factor. This SMR-first pipeline, which additionally required a suggestive SMR association and a non-significant HEIDI test, retained three proteins that also colocalized with POAG (PP4 > 0.5): LILRB5, TMEM190 and RGMB (Supplementary Table S9). The unbiased colocalization scan above additionally surfaced LILRB2, which was not among the proteins passing the SMR/HEIDI filter, so the two approaches are concordant for LILRB5, TMEM190 and RGMB. To make candidate prioritization explicit, RGMB was carried forward under a pre-specified hierarchy: among proteins passing the SMR, HEIDI and colocalization filters in the East Asian arm, we prioritized those additionally showing (i) evidence consistent with an IOP-independent mechanism and (ii) a biologically plausible link to retinal ganglion-cell vulnerability. LILRB2 was not among the proteins passing the SMR/HEIDI filter, and LILRB5, although it passed, rested on a five-variant HEIDI test, could not be separated from the neighbouring LILRB2 signal, and lacked a mechanistic link to retinal ganglion cells; TMEM190, although it passed the filters, behaved in an outcome-GWAS-dependent manner (see Discussion) and lacked a comparable mechanistic rationale; RGMB met all of these criteria and was therefore retained as the prioritized primary candidate rather than an established factor.

**Figure 2.**
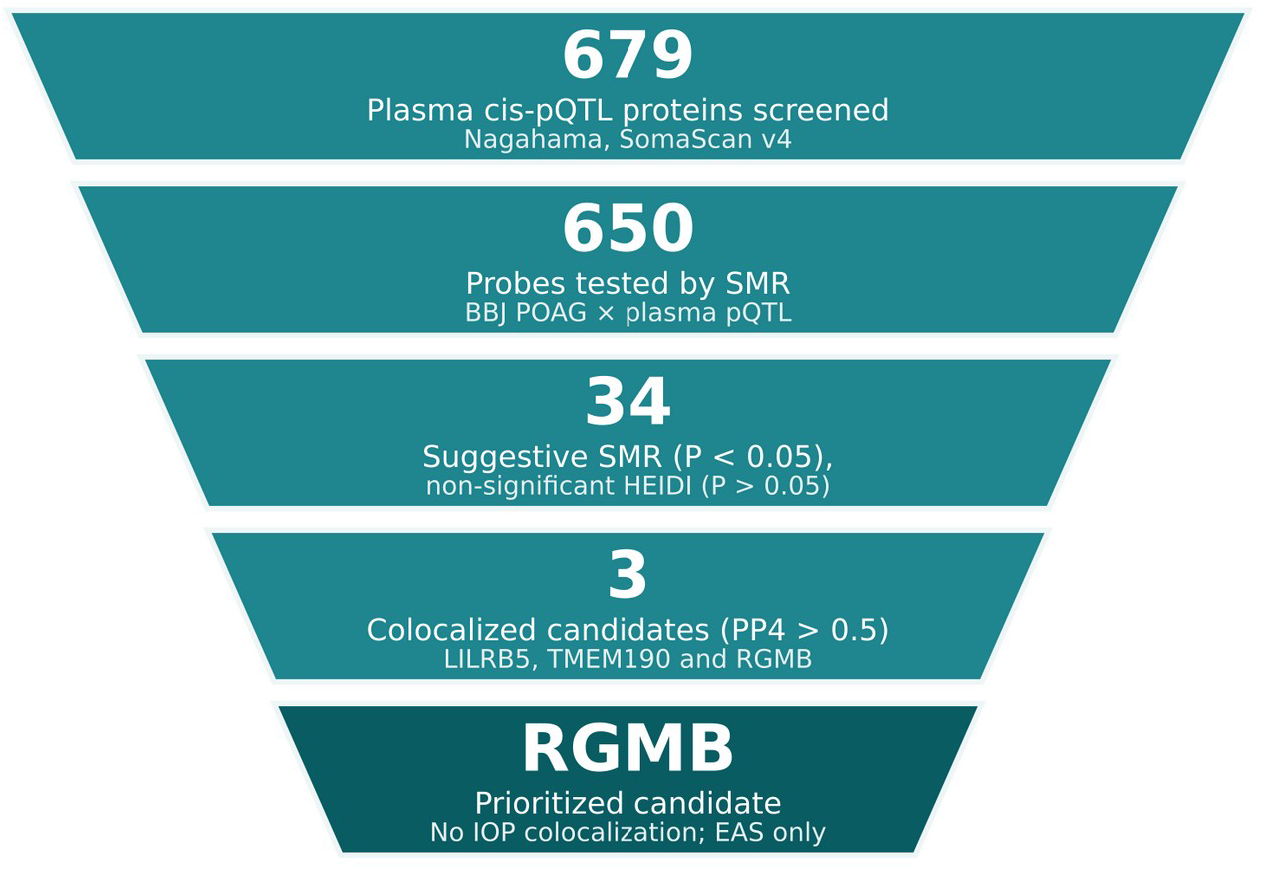
Data-driven prioritization strategy for candidate causal proteins. Plasma proteins with available *cis*-pQTL instruments in the Nagahama resource (679) were screened for a shared genetic signal with BBJ POAG by SMR (650 probes tested), filtered to suggestive associations (SMR *P* < 0.05 with non-significant HEIDI *P* > 0.05; 34 candidates), and narrowed by Bayesian colocalization (PP4 > 0.5) to three proteins (LILRB5, TMEM190 and RGMB). Although no protein survived proteome-wide (Bonferroni) correction, RGMB was carried forward as the most interpretable candidate because it showed concordant evidence across SMR, HEIDI, colocalization, and biological-plausibility analyses; LILRB5 was not prioritized because its HEIDI test rested on five variants and its colocalization could not be separated from that of the neighbouring LILRB2. RGMB was prioritized as a candidate that did not colocalize with IOP and was detected only in the East Asian data. No protein survived Bonferroni correction; the strategy is exploratory and hypothesis-generating.

### The RGMB locus does not colocalize with IOP

To assess whether the RGMB signal acts through elevated IOP, we colocalized RGMB pQTLs with two European IOP datasets. In IGGC (*n* = 31,269), there was no evidence of colocalization with IOP (PP4 = 0.007; Figure 3), with the posterior overwhelmingly favoring a signal in only one dataset (PP.H2 = 0.977). The same pattern was observed in the larger Pan-UKBB European IOP GWAS (*n* = 87,199; PP4 = 0.003; PP.H2 = 0.988). Consistent with these results, the RGMB region was not among the 21 genome-wide significant loci of the Japanese TMM IOP GWAS (*n* = 33,452). Across both European cohorts the absence of colocalization reflected a lack of detectable IOP signal in the RGMB window rather than a competing, distinct signal.

**Figure 3.**
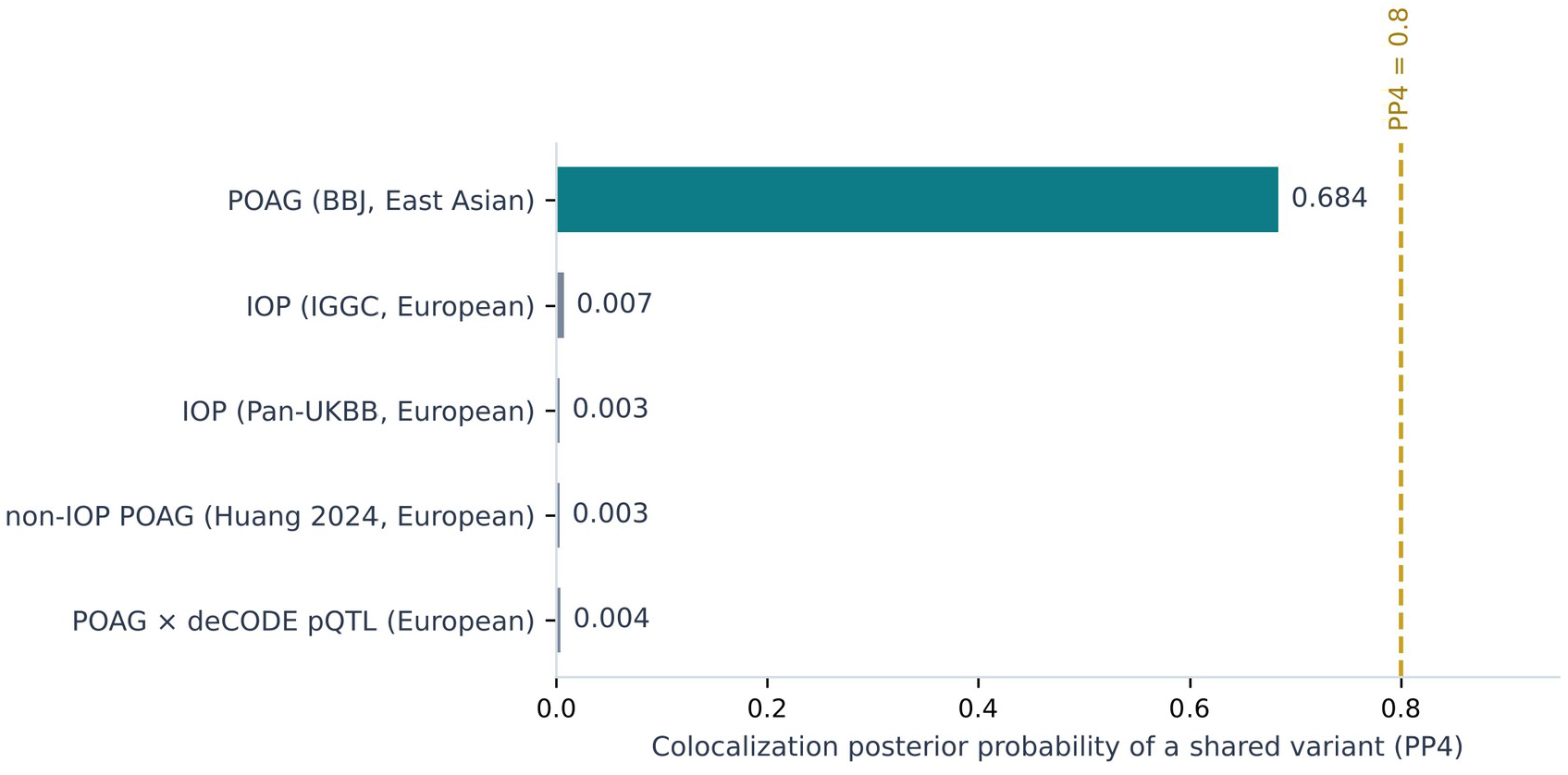
Colocalization of plasma RGMB across POAG and IOP datasets. Posterior probability of a shared causal variant (PP4) between the plasma-RGMB *cis*-pQTL and each outcome. Suggestive colocalization was observed only with East Asian POAG (BBJ; PP4 = 0.684), whereas European IOP (IGGC and Pan-UKBB), the European IOP-independent POAG component^6^, and a fully European same-platform analysis (deCODE pQTL × Gharahkhani POAG) all showed no colocalization. The dashed line marks the conventional PP4 = 0.8 threshold for strong colocalization. PP4 is presented as a continuous measure of colocalization evidence; candidate prioritization in this study used PP4 > 0.5 in combination with SMR and HEIDI evidence, whereas PP4 ≥ 0.8 denotes strong colocalization.

### No colocalization with a European IOP-independent POAG component

To test the IOP-independent hypothesis more directly, we colocalized RGMB pQTLs with the non-IOP POAG latent component of Huang et al.^6^, generated by Genomic SEM from European POAG and IOP inputs. RGMB did not colocalize with this European-derived component (PP4 = 0.003; PP.H2 = 0.986). Thus, although the RGMB–POAG colocalization was detectable in the East Asian (BBJ) data, the RGMB region showed no signal across three independent European comparators (two IOP datasets and the non-IOP POAG component), and RGMB is not reported among genome-wide significant European POAG loci. Taken together, these results are consistent with an IOP-independent locus whose detectable effect may reflect an ancestry-enriched effect in East Asian populations. A non-significant HEIDI test reduces, but does not exclude, horizontal pleiotropy as an alternative explanation for the SMR association. We interpret this as a hypothesis rather than an established fact and, to probe whether the European nulls could be technical artifacts, performed a within-European analysis and an allele-frequency comparison, described next.

### A within-European analysis also shows no colocalization, and the lead variant is common in both ancestries

To assess whether the European null results might reflect a mismatch in ancestry between the exposure (East Asian pQTL) and outcome (European GWAS), or the proteomic platform, rather than a difference in the underlying genetics, we performed two additional checks. First, we repeated the colocalization entirely within European data, using a European RGMB *cis*-pQTL from deCODE (SomaScan v4; the same aptamer, SeqId 3331-8, *n* = 35,559) and the European POAG meta-analysis of Gharahkhani et al.^15^ (GCST90011766; 16,677 cases and 199,580 controls). Across the RGMB *cis*-window (9,682 shared variants), RGMB did not colocalize with European POAG (PP4 = 0.004), with the posterior divided between distinct causal variants (PP.H3 = 0.381) and a signal in only one dataset (PP.H2 = 0.614). Because both exposure and outcome were European and the same proteomic platform was used, this null cannot be attributed to cross-ancestry exposure–outcome mismatch or to platform differences. Second, the lead variant rs1037599 (effect allele G) is common in both ancestries (G-allele frequency ≈ 0.35 in the East Asian pQTL data and ≈ 0.47 in European data), so the absence of a European signal is unlikely to reflect low allele frequency or limited power. Together, these analyses make differences in LD structure or a genuinely ancestry-enriched genetic effect a more likely explanation for the East Asian–detected signal than these technical alternatives, although they do not by themselves establish an ancestry-specific causal effect.

### A formal test of cross-ancestry heterogeneity

The classification of proteins by whether PP4 exceeds 0.5 is a convenience threshold and not a test of ancestry heterogeneity. We therefore compared the per-allele POAG effect of rs1037599 between ancestries directly. The two estimates differed significantly (East Asian β = +0.083, SE 0.026, P = 1.5 × 10⁻s; European β = −0.0016, SE 0.0129, P = 0.90; Cochran’s Q = 8.50, df = 1, P = 0.0036; I² = 88.2%), and heterogeneity was significant at all 22 East Asian LD proxies of the lead variant (median I² = 87.5%; Benjamini–Hochberg FDR < 0.01 for every proxy), including the regulatory candidate rs1563318 (Q = 7.56, P = 0.006; Supplementary Table S7). Importantly, the asymmetry in sample size favours the European dataset on both sides of the analysis: the European POAG meta-analysis has 4.2 times as many cases and a standard error half as large (0.0129 versus 0.0260), and the European cis-pQTL instrument is the more precisely estimated of the two (z = 15.1 versus 8.4). The European 95% confidence interval (−0.027 to +0.024) excludes the East Asian point estimate, and an effect of the East Asian magnitude would have been detected in the European data at Z = 6.4 (P = 1.2 × 10⁻¹⁰). The absence of a European association is therefore not attributable to limited power. We note, however, that rs1037599 was prioritized on the basis of its colocalization in the East Asian data, so the East Asian effect estimate may be subject to selection bias; the heterogeneity test should accordingly be read as a quantification of the observed difference rather than as independent confirmation of an ancestry-specific effect.

### Cross-ancestry fine-mapping points to an ancestry-differential effect rather than to LD divergence

In the East Asian plasma pQTL data, fine-mapping of the RGMB *cis*-region resolved a single 95% credible set (16 variants) whose top variant by PIP was the POAG lead SNP rs1037599 (PIP = 0.26). The variant most likely to regulate plasma RGMB in East Asians therefore coincided with the POAG association signal, providing a mechanistic correlate of the colocalization (PP4 = 0.684). In the better-powered European deCODE data (*n* = 35,559), the same region resolved into ten independent credible sets; the dominant signal was a distinct variant, rs62369889 (PIP = 1.00), and rs1037599 was not contained in any European credible set, appearing only as a secondary signal.

Cross-ancestry comparison showed that the rs1037599 block itself is conserved in both ancestries: its closest proxies were in complete LD in both panels (r² = 1.0), and the wider block remained strongly correlated in Europeans (core-block r² ≈ 0.84 in East Asians versus ≈ 0.75 in Europeans). A simple explanation in which the lead variant fails to tag the causal variant in Europeans (LD-untagging) is therefore not supported for the immediate block; appreciable LD divergence was confined to secondary blocks (for example, the European rs62369959 cluster, r² = 0.35 in East Asians versus 0.10 in Europeans).

Critically, although the rs1037599 block is conserved in European LD and demonstrably regulates plasma RGMB in Europeans (a strong deCODE *cis*-pQTL), none of these variants showed any association with POAG in Europeans: rs1037599 itself (β = −0.002, *P* = 0.90), the 20-variant block (minimum *P* = 0.51, median 0.70; 0/20 with *P* < 0.05), and the European-dominant regulator rs62369889 (*P* = 0.43) were all null, and across all 59 RGMB *cis*-variants examined none approached genome-wide significance (minimum *P* = 0.046). Fine-mapping of the East Asian POAG GWAS over the same region did not converge to a credible set (lead *z* = 3.19; maximum PIP = 0.03), consistent with the limited power of the BBJ POAG GWAS (3,980 cases). Taken together with the exclusion of allele-frequency, platform, and exposure–outcome ancestry artifacts described above, these analyses indicate that the absence of European colocalization is most consistent with, but not definitive for, an ancestry-differential relationship between RGMB regulation and POAG risk — that is, RGMB *cis*-regulatory variation that is shared across ancestries but associated with POAG risk only in East Asians — rather than with simple LD divergence. This remains a candidate interpretation: it rests on a single East Asian discovery without an independent public East Asian replication dataset, the colocalization evidence is correlational and did not survive Bonferroni correction, and the underlying causal variant could not be resolved (Figure 4).

**Figure 4.**
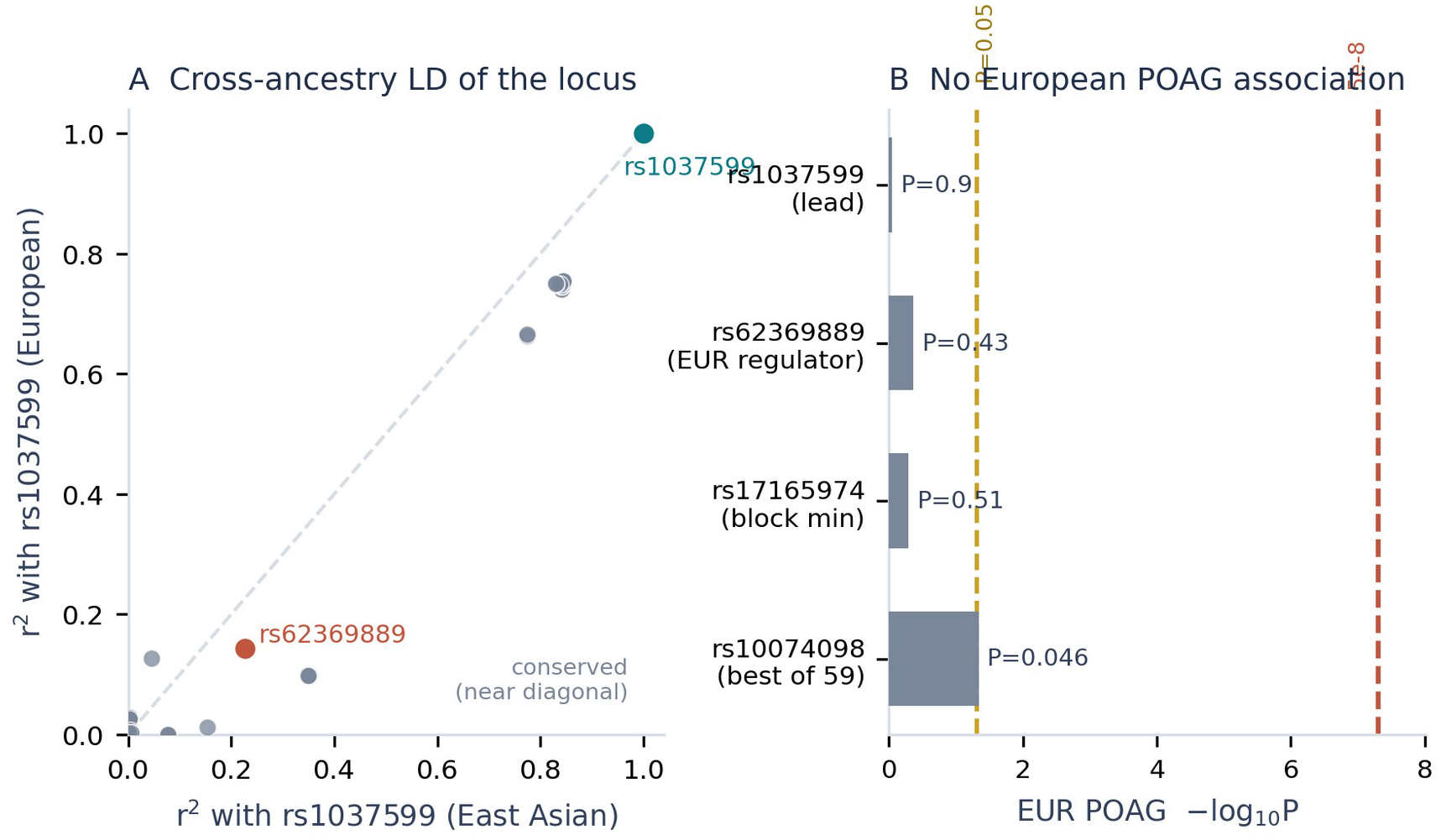
Cross-ancestry fine-mapping at the RGMB locus. (A) Squared correlation (r²) of each cis variant with the POAG lead rs1037599 in East Asian (x-axis) versus European (y-axis) 1000 Genomes reference panels. Variants near the diagonal are conserved across ancestries; the immediate rs1037599 block (upper right) is conserved, whereas the European-dominant RGMB regulator rs62369889 and a secondary cluster are tagged less in Europeans. (B) European POAG association (−log₁₀P) for the lead variant, the European-dominant regulator, the strongest variant in the rs1037599 block, and the strongest of all 59 cis variants examined. The strongest of the 59 cis variants reaches nominal significance (P = 0.046; dashed line at P = 0.05), but no variant approaches the genome-wide threshold (P = 5 × 10⁻⁸), and neither rs1037599 (P = 0.90) nor any variant in its LD block (minimum P = 0.51) is nominally significant.

### Multi-signal colocalization supports the East Asian signal and the European null

Because coloc.abf assumes a single causal variant per trait and the European pQTL region resolved into ten independent credible sets, we repeated the RGMB colocalization with methods that accommodate multiple signals. After removal of variants inconsistent with the reference LD (22 of 924 variants in the East Asian arm and 13 of 937 in the European arm), the East Asian pQTL region resolved into a single 95% credible set in every window and regularization setting examined, with rs1037599 as the top variant (posterior inclusion probability 0.27–0.59; credible-set purity 0.84– 0.91); the single-causal-variant assumption is therefore appropriate for the East Asian arm (Supplementary Table S5). The European region resolved into five to ten credible sets depending on window width, reproducing the ten sets reported above at ±500 kb. Consistent with the fine-mapping, masking and conditioning each returned a single signal pair in East Asians, led by rs1037599, with posteriors close to the primary analysis (PP4 = 0.691 by masking and 0.350 by conditioning, versus 0.692 by coloc.abf over the same 902 variants; the lower conditional estimate was offset by PP.H1 rather than by PP.H3, that is, by the weakness of the outcome signal rather than by evidence for distinct causal variants, PP.H3 = 0.027). In Europeans, masking identified three distinct signal pairs and conditioning six, and none colocalized with POAG (masking PP4 = 0.012, 0.007 and 0.017; conditioning PP4 ≤ 2.4 × 10⁻⁵ with PP.H3 ≥ 0.95, that is, positively favouring distinct causal variants rather than merely failing to support a shared one). Across the sensitivity analyses the European posteriors remained below 0.02 except for a single masked signal pair at ±150 kb (PP4 = 0.19; Supplementary Table S6). The European null is therefore not an artefact of collapsing multiple independent pQTL signals into a single-variant model, and the East Asian result is not inflated by it.

### Retinal single-cell context of the RGM–neogenin–BMP axis

To relate the circulating-protein findings to ocular tissue, we examined the CZ CELLxGENE Discover human retina atlas. Atlas labeling and scaling were validated by the expected RGC-selective expression of RBPMS (∼85% of RGCs). RGMB transcript abundance was low and broadly distributed across retinal cell types (∼0.41% of RGCs; scaled expression ≈ 0.19), which is consistent with the low steady-state transcription expected for a low-abundance membrane co-receptor and with droplet-based dropout. Low transcript abundance does not exclude functional significance for membrane-bound signaling receptors such as RGMB, whose activity depends on cell-surface availability rather than high steady-state mRNA levels. In contrast, the RGMB receptor NEO1 and the BMP type-I receptor BMPR1A were expressed across retinal neurons, including RGCs, rods, and bipolar cells (for example, BMPR1A in ∼41% of rods and ∼12% of RGCs; NEO1 in ∼27% of rods and ∼9% of RGCs). This co-expression pattern is consistent with the presence of an intact RGM–neogenin–BMP receptor architecture in human retina, but expression alone does not establish pathway activity or a functional role in disease (Figure 5).

**Figure 5.**
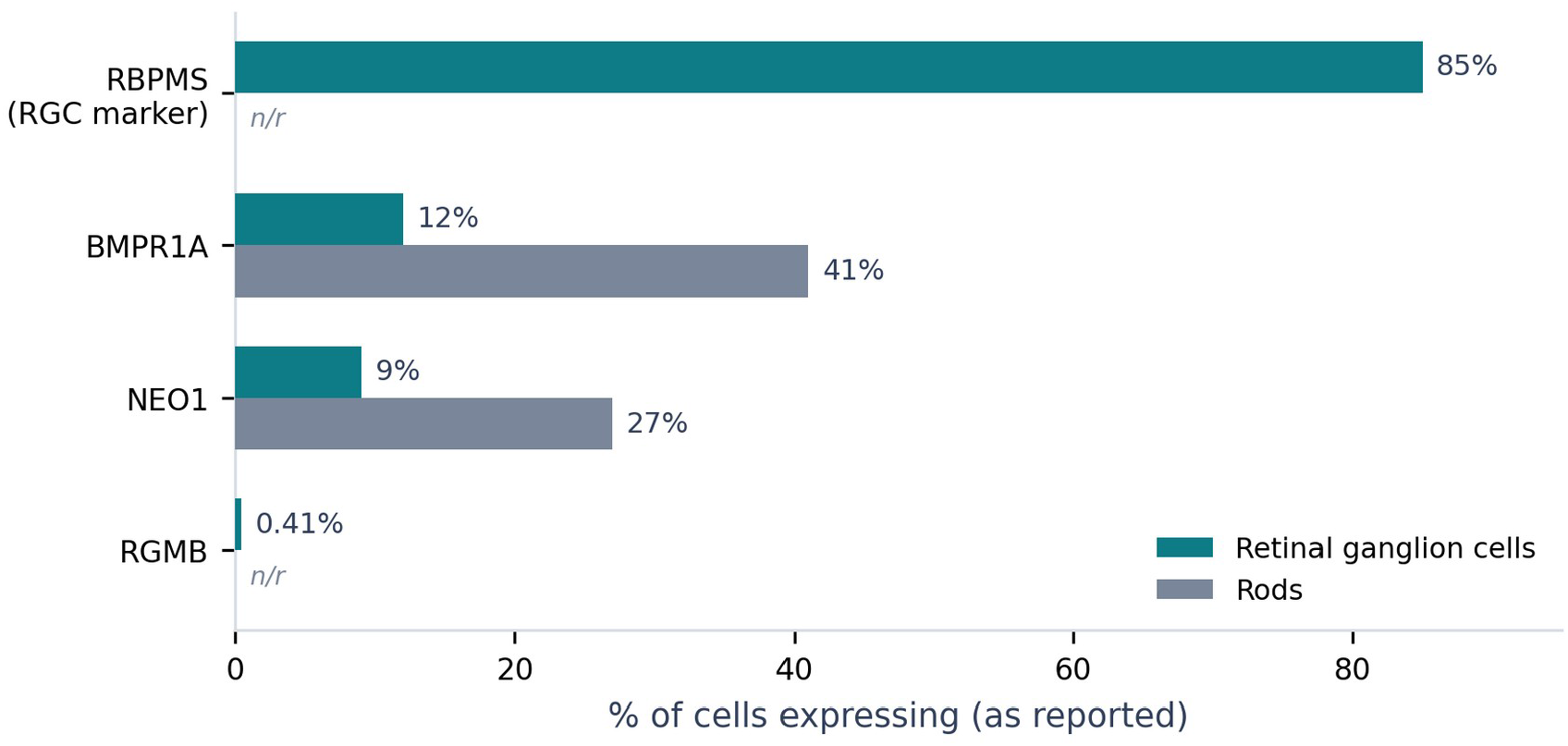
Retinal single-cell expression of RGMB-pathway components. Percentage of cells expressing each gene in retinal ganglion cells and rods in the CZ CELLxGENE Discover human retina atlas, for the values reported in the text. The RGC marker RBPMS confirms atlas labeling and scaling; RGMB transcript is sparse, whereas its receptor NEO1 and the BMP type-I receptor BMPR1A are expressed across retinal neurons. n/r, not reported.

### RGMB and its receptors are expressed in trabecular meshwork cells but do not respond to TGF-β2

Because the conventional outflow pathway is central to POAG pathophysiology, and because plasma protein levels need not reflect ocular tissue, we examined RGMB-pathway expression in primary human trabecular meshwork (TM) cells. In GSE236302, RGMB was detected in all 15 samples at 22 CPM, close to the median of the expressed transcriptome (52nd percentile among 15,973 genes with mean CPM > 1) and above TGFB2 itself (15 CPM, 44th percentile); NEO1 (28 CPM), BMPR1A (71 CPM) and BMPR2 (85 CPM) were also expressed (57th, 79th and 83rd percentiles). TM identity was supported by high MYOC expression (98.5th percentile). RGMB was likewise detected in all eight samples of the independent dataset GSE195756 (25–96 CPM), as were NEO1, BMPR1A and BMPR2. The low retinal transcript abundance of RGMB therefore does not extend to the trabecular meshwork, where the ligand and its receptor are present at ordinary levels (Supplementary Table S8).

TGF-β2 treatment of the same TM cells produced the expected fibrotic response (SERPINE1 log2 fold change = +1.89, P = 2.5 × 10⁻⁴; ACTA2 +1.24; COL1A1 +1.12; FN1 +0.86), yet RGMB and NEO1 were unchanged (log2 fold change = −0.001, P = 0.997 and −0.04, P = 0.81), as was BMPR1A (−0.04, P = 0.68). BMP ligands did shift (BMP2 +1.50 and BMP4 −1.45, both FDR < 0.01), consistent with the reported antagonism between BMP-4 and TGF-β2 in the trabecular meshworks⁰. RGMB and its receptor are thus present in TM cells but sit outside the TGF-β2–driven fibrotic programme that is most closely tied to outflow resistance, which is consistent with, although it does not establish, the IOP-independent interpretation of the genetic findings. These data derive from cultured TM cells rather than native tissue and are transcriptomic, so they speak to the presence of the pathway and its response to a single stimulus rather than to its activity in vivo.

## Discussion

Using a data-driven proteogenomic framework — SMR/HEIDI, multi-dataset colocalization, comparison with a non-IOP POAG component, and single-cell retinal mapping — we prioritized plasma RGMB as a candidate IOP-independent risk factor for POAG. The most consistent feature of the data is that the RGMB association was detectable in East Asian POAG but absent across all European comparators, while showing no colocalization with IOP in either of two European cohorts or with the Japanese TMM IOP loci. We therefore interpret RGMB as a candidate pressure-independent locus, pending further validation, rather than a confirmed causal factor.

This prioritization is reinforced by a symmetric, proteome-wide comparison of colocalization profiles between ancestries. Using the same platform and method, the proteins colocalizing with POAG in East Asians (LILRB2, LILRB5, TMEM190 and RGMB) and in Europeans (ITIH1, ADPRHL2, PLXNA1, SVEP1 and MMAB) formed non-overlapping sets, with no protein colocalizing in both; given the moderate posterior probabilities, this non-overlap should be interpreted cautiously (Figure 1; Tables 1 and 2). RGMB was the most biologically coherent of the East Asian–enriched proteins for an IOP-independent mechanism, whereas the European–enriched set included genes with prior glaucoma or ocular relevance (for example SVEP1 and PLXNA1), indicating that the scan recovered credible signals on both sides. This profile-level divergence should be interpreted cautiously: no protein reached strong colocalization (PP4 > 0.8) in Europeans, so the absence of shared signals may reflect ancestry-differential effects, LD differences, or limited power rather than a definitive biological dichotomy, and the immune receptors LILRB2 and LILRB5 lie in the LD-complex chromosome-19 LILR cluster and rest on fewer shared variants. Their higher PP4 relative to RGMB, together with near-identical values for the two homologous, closely linked genes, is most consistent with a shared regional signal or multiple/linked causal variants rather than a single shared variant, and only LILRB5 passed the SMR/HEIDI filter, on a five-variant HEIDI test; RGMB, supported concordantly across SMR, HEIDI and colocalization, is the more interpretable candidate.

Our European-enriched signals are partly concordant with prior European-only proteome-wide analyses of POAG, with this concordance carried mainly by SVEP1. SVEP1, our strongest European-enriched protein, was among the plasma proteins nominated as POAG drug targets using deCODE pQTL in a prior European analysis^8^ and is an established Schlemm’s-canal/IOP-related modifier. The remaining four European-enriched proteins were not nominated as colocalized targets in those studies: notably, the inter-α-trypsin inhibitor heavy chain recovered here was ITIH1, whereas the prior tier-one signal was the paralogous but distinct ITIH3^7^, and ADPRHL2, PLXNA1 and MMAB did not appear among the prior nominated targets. Because those analyses used different POAG outcome GWAS than the Gharahkhani meta-analysis used here, this partial non-overlap is consistent with sensitivity to the choice of outcome GWAS (as illustrated for TMEM190 below) rather than a contradiction between studies. TMEM190 is an instructive exception: it showed outcome-GWAS–dependent colocalization (detected in a European discovery cohort using Global Biobank Meta-analysis Initiative and FinnGen POAG, but not in our deCODE × Gharahkhani comparison) and lacked a comparable mechanistic rationale for an IOP-independent retinal ganglion-cell pathway; therefore, although it passed the statistical filters, it was not prioritized further. This underscores caution when interpreting ancestry-enriched classifications based on moderate posterior probabilities. Reassuringly, the estimated direction of effect was concordant across ancestries: higher plasma TMEM190 was associated with lower POAG risk both in our East Asian analysis (*b*_SMR_ = −0.086) and in that European study (*b*_SMR_ = −0.049 and −0.040 in its discovery and replication cohorts, respectively), and our East Asian lead variant (rs4806666) matched that study’s replication-cohort lead variant. The cross-ancestry difference therefore concerns whether the signal reaches the colocalization threshold under a given outcome GWAS, not the direction of the TMEM190–POAG relationship. Importantly, RGMB itself was not nominated in those European analyses; the related axon-guidance molecule RGMA appeared only as weak (suggestive, non-colocalizing) European evidence. RGMA was not assayed in the Nagahama panel and so could not be evaluated in either arm of our symmetric scan; the East Asian–enriched signal we report is therefore specific to RGMB rather than to the RGM family in general (RGMB and RGMA are paralogues encoded on different chromosomes, 5q15 and 15q26), and the RGMB-versus-RGMA distinction should be kept in mind when comparing across studies. To make this comparison explicit, and to separate genuine cross-ancestry differences from gaps in panel coverage, we cross-tabulated every protein nominated by the two prior European studies against its status in the present scan (Supplementary Figure S3; Supplementary Table S1). Of the nine prior-European targets, five (ENPP5, NCR1, TIMP3, SERPINF1 and OXT) were assayed in the Nagahama panel yet did not colocalize in either ancestry arm (PP4 < 0.5), indicating a genuine difference attributable to the outcome GWAS and method rather than to coverage, whereas four — including the tier-one targets ROBO1, FOXO3 and ITIH3 — were not represented among the 679 aptamers and were therefore not evaluable here. The method-level evidence for RGMB (SMR, HEIDI and colocalization) across the two arms and the overall concordance with the prior European studies are summarized in Supplementary Figures S1 and S2. This resolution indicates that the non-overlap with European targets is driven partly by genuine outcome-dependent differences and partly by panel coverage, rather than by a contradiction between studies.

The contrast between the East Asian and European results is consistent with — but does not by itself establish — an East Asian–enriched effect. Several technical explanations can now be addressed directly. The European nulls are not readily explained by exposure–outcome ancestry mismatch or by proteomic platform, because a fully European, same-platform analysis (deCODE RGMB pQTL × European POAG) also showed no colocalization; nor by allele frequency or statistical power, because the lead variant is common in both ancestries. The remaining explanations are differences in LD structure between ancestries or a genuinely ancestry-enriched genetic effect.

Cross-ancestry fine-mapping helps adjudicate between these. The immediate rs1037599 block is conserved in European LD (core-block r² ≈ 0.75 versus ≈ 0.84 in East Asians) and demonstrably regulates plasma RGMB in Europeans, yet shows no association with European POAG (block minimum *P* = 0.51); this argues against a simple failure of the lead variant to tag the causal site in Europeans (LD-untagging) and favours an ancestry-differential relationship between RGMB regulation and POAG risk. The genomic position of the lead variant is consistent with a regulatory framing: rs1037599 lies approximately 320 kb upstream (5′) of the RGMB transcription start site (chr5:98,447,787 versus the RGMB gene at chr5:98,768,004–98,798,643, GRCh38; Figure 6), consistent with a distal regulatory variant — or a tag for one — rather than a coding change. Exploratory regulatory annotation (Supplementary Note; Supplementary Figures S4 and S5) indicated that rs1037599 itself overlaps no candidate cis-regulatory element (RegulomeDB rank 6) but tags an LD proxy, rs1563318 (r² = 0.84), located within a distal enhancer-like element with strong regulatory evidence (rank 1b), consistent with the lead variant tagging a regulatory module rather than acting directly. Consistent with this, both variants were significant *cis*-eQTLs for RGMB in GTEx v10: across the tissues tested, the plasma-RGMB-raising allele (G) was associated with higher RGMB mRNA — concordantly for rs1563318 (positive in all nine significant tissues; e.g., gastroesophageal junction NES = +0.26, *P* = 1.4 × 10⁻¹⁸) and in six of seven significant tissues for rs1037599 — in the same direction as the plasma pQTL, although no ocular tissue is represented in GTEx. Notably, rs1563318—the variant singled out by chromatin annotation (RegulomeDB rank 1b, within the enhancer-like cCRE)—is also the cleaner expression signal, an eQTL concordant in all nine significant tissues versus six of seven for the lead, so the chromatin and expression evidence converge on the same element, even though tight LD prevents either from formally discriminating it from rs1037599. That the RGMB-raising allele raises both RGMB mRNA and plasma RGMB in the same direction further points to a transcriptional, *cis*-regulatory mechanism rather than a post-transcriptional one. These inferences rest, however, on non-ocular tissues, and the physical enhancer–promoter contact (∼320 kb) and the responsible transcription factor remain untested, so the causal element, variant, tissue, and effector await ancestry– and tissue-matched functional validation. We nonetheless cannot formally distinguish an ancestry-specific causal effect from residual differences in regulatory architecture (that is, which *cis* signal dominates in each ancestry). Establishing an ancestry-specific causal effect would require independent East Asian POAG cohorts and, where available, ancestry-matched ocular molecular data; notably, no independent East Asian POAG summary-statistic dataset distinct from the discovery sample is currently available in the public domain, which constrains replication.

**Figure 6.**
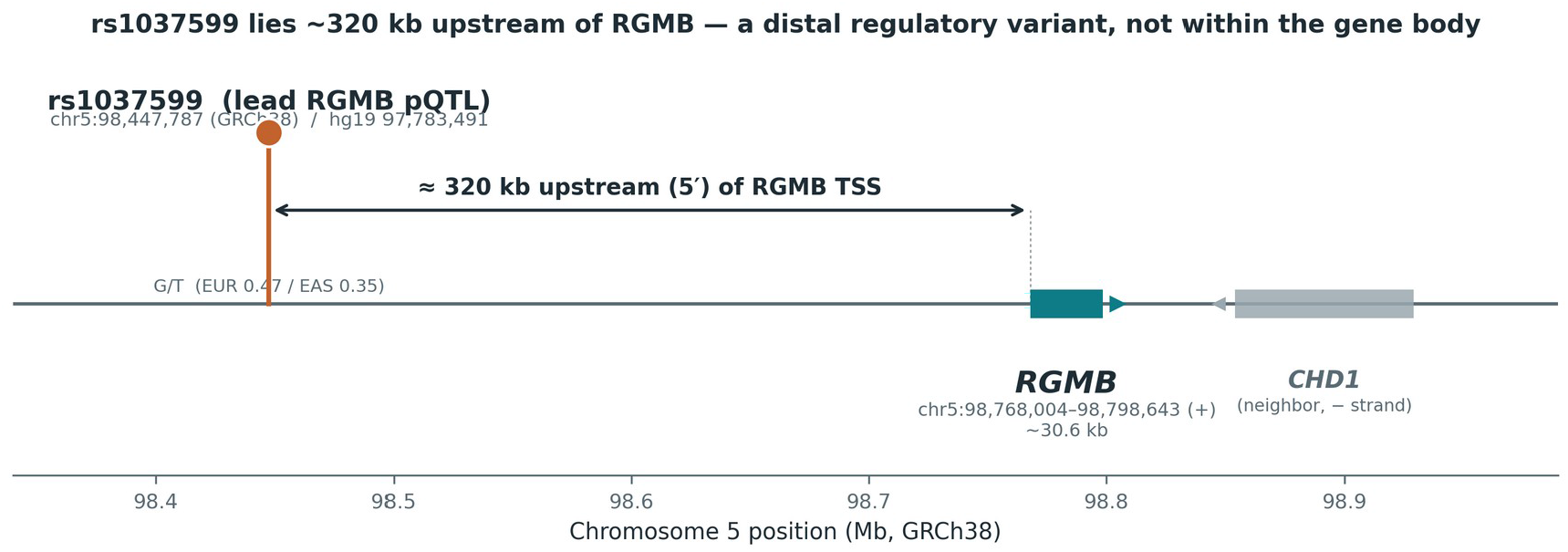
Genomic context of the lead RGMB pQTL variant rs1037599 (chromosome 5, GRCh38). The lead plasma-RGMB pQTL variant rs1037599 (chr5:98,447,787; effect allele G, frequency ≈0.47 in Europeans and ≈0.35 in East Asians) is located approximately 320 kb upstream (5′) of the RGMB gene body (chr5:98,768,004–98,798,643, plus strand). The neighbouring gene CHD1 (minus strand) is shown for context. The position indicates that rs1037599 is a distal upstream variant relative to RGMB rather than a within-gene variant.

These genetic observations align in direction with the longstanding epidemiological pattern in East Asia, where NTG predominates and conventional IOP-lowering therapy often fails to halt progression. An IOP-independent, potentially ancestry-enriched candidate such as RGMB is of interest in this context, but the present analyses are correlational and do not demonstrate that RGMB causally contributes to RGC loss or that it underlies the NTG phenotype.

RGMB is a member of the repulsive guidance molecule family and functions as a BMP co-receptor acting through neogenin (NEO1) to influence axon guidance, cell survival, and structural plasticity. Within the central nervous system, the related molecule RGMa is upregulated after neuronal injury and can inhibit axonal regeneration. In the eye specifically, RGMa promotes retinal ganglion cell survival in vitro and in vivo through neogenin^28^, whereas antibody blockade of RGMa protects the retinal nerve fiber layer and promotes optic nerve axon regeneration after injury^29^, placing the RGM–neogenin axis directly within retinal ganglion cell survival and axon biology. These known roles offer a biologically plausible link to RGC vulnerability, and our single-cell results indicate that the receptor components of this pathway are present in human retinal neurons. As a BMP co-receptor, RGMB could in principle also intersect canonical POAG pathophysiology: bone morphogenetic proteins, their receptors, and the antagonist gremlin are expressed in the trabecular meshwork and optic nerve head, where the BMP–TGF-β2 axis governs extracellular matrix turnover, aqueous outflow resistance, and intraocular pressure^30^. RGMB might therefore act through an IOP-independent neuronal route, an IOP-related anterior-segment route, or both; and because RGM proteins can either potentiate or restrain BMP signaling depending on ligand and tissue context, even the direction of any RGMB effect cannot be inferred from expression alone. However, the retinal data are transcriptomic and derived from non-diseased tissue, the plasma pQTL reflects circulating rather than ocular protein, and the direction and mechanism of any effect on RGC survival remain to be tested experimentally.

### Independent instrument validation of the RGMB cis-pQTL in an East Asian cohort

To address the reliance on a single East Asian discovery sample, we sought independent confirmation of the RGMB cis-pQTL in a separate East Asian cohort assayed on a different proteomic platform. In the China Kadoorie Biobank (CKB; n ≈ 3,974; Olink Explore antibody-based assay), the discovery lead variant rs1037599 was a genome-wide-significant cis-pQTL for RGMB (β = +0.18 per G allele; P = 2.9 × 10⁻¹⁴; G-allele frequency 0.39), and the same G allele raised plasma RGMB as in the Nagahama discovery data (SomaScan v4 aptamer assay; β = +0.30 per G allele; P = 9.8 × 10⁻¹⁷). Because the two cohorts differ in ancestry (Japanese versus Chinese) and in proteomic technology (aptamer versus antibody), this concordance argues against an aptamer-specific artifact and supports a genuine protein-level cis effect; effect-size magnitudes are platform-dependent and were not compared, only the effect allele and the sign.

Extending the comparison to Europeans, the same cis-pQTL was directionally consistent in deCODE (SomaScan v4; β = +0.12 per G allele; P ≈ 2 × 10⁻⁵¹), so the G allele of rs1037599 raises plasma RGMB across all three ancestries examined and both platforms (Table 3). The shared instrument direction and broadly similar allele frequencies (0.35–0.47) indicate that the East Asian–enriched colocalization with POAG is not attributable to ancestry differences in the instrument itself — the European instrument is in fact the most precisely estimated — but rather to the relationship between RGMB and POAG, which colocalized in East Asians (PP4 = 0.684) and not in Europeans (PP4 = 0.004).

**Table 3.** Direction of the RGMB rs1037599 cis-pQTL across three ancestries and two proteomic platforms.

| Cohort<br>(ancestry) | Platform | Effect<br>allele | β (plasma<br>RGMB) | G-allele<br>frequency | cis-pQTL P |
| --- | --- | --- | --- | --- | --- |
| Nagahama<br>(Japanese) | SomaScan<br>v4 | G | +0.30 | 0.35 | 9.8 × 10<sup>-17</sup> |
| CKB<br>(Chinese) | Olink Explore | G | +0.18 | 0.39 | 2.9 × 10<sup>-14</sup> |
| deCODE<br>(European) | SomaScan<br>v4 | G | +0.12 | 0.47 | 2 × 10<sup>-51</sup> |

β values are on platform-specific scales (SomaScan, relative fluorescence units; Olink, normalized protein expression) and are not directly comparable in magnitude; only the effect allele and the sign of the effect are compared across cohorts. CKB, China Kadoorie Biobank.

We additionally repeated the colocalization using the independent CKB cis-pQTL as the instrument against the same East Asian POAG GWAS (976 shared variants). Under coloc default priors this yielded a posterior probability of a shared causal variant of PP4 = 0.73, consistent with the discovery estimate (0.684). This support was sensitive to the prior probability of shared causality (p12 = 1 × 10⁻⁵, 1 × 10⁻⁶, and 1 × 10⁻⁷ gave PP4 = 0.73, 0.22, and 0.03, respectively; full posteriors in Supplementary Table S2), reflecting the limited power of the East Asian POAG signal at this locus rather than the strength of the pQTL. This analysis therefore validates the instrument — an independent East Asian cis-pQTL — rather than replicating an independent outcome GWAS, because the outcome dataset is the same as in the discovery analysis; an adequately powered, independent East Asian POAG GWAS, which is not currently available in the public domain, will be required for outcome-level replication. The direction was concordant on the outcome side, with the RGMB-raising G allele associated with higher POAG risk in Biobank Japan (log-odds ratio = +0.083).

## Limitations

Several limitations temper interpretation. First, no protein survived Bonferroni correction, so the prioritization is exploratory and requires replication. Second, the RGMB–POAG colocalization (PP4 = 0.684) is suggestive rather than strong (conventional threshold ≈ 0.8). Third, although a within-European same-platform analysis and the common frequency of the lead variant argue against proteomic-platform, allele-frequency, and power artifacts, the East Asian–enriched interpretation still rests on a single East Asian discovery sample; no independent East Asian POAG summary-statistic dataset is currently available for replication, and differences in LD structure versus a genuine ancestry-specific effect cannot be distinguished with the present data. Fourth, SMR assumes that the instrument acts on the outcome through the exposure and can be biased by horizontal pleiotropy despite a non-significant HEIDI test, which assesses linkage but does not by itself exclude horizontal pleiotropy. Fifth, the single-cell evidence is transcriptomic, from healthy donors, and subject to dropout, and therefore cannot confirm protein-level activity or disease relevance. Sixth, no ancestry-matched ocular (retinal) eQTL resource is currently available for East Asian populations — existing retinal eQTL datasets are of European ancestry — which currently precludes a tissue– and ancestry-matched expression colocalization. Seventh, fine-mapping of the East Asian POAG GWAS did not converge to a credible set, reflecting the limited power of the discovery sample; the mechanistic link between the POAG signal and RGMB regulation therefore rests on colocalization and pQTL fine-mapping rather than on overlapping GWAS credible sets, and LD for fine-mapping was estimated from reference panels of approximately 500 individuals rather than in-sample. Eighth, the null

European POAG look-up does not exclude a small effect below current detection limits. Ninth, the use of summary-level data precludes assessment of interaction or non-linear effects. Tenth, in the cross-ancestry profile comparison no protein reached strong colocalization (PP4 > 0.8) in Europeans, so the non-overlap of ancestry profiles — including the East Asian–enriched classification of RGMB — reflects moderate posterior probabilities and may be influenced by differences in LD or power; reassuringly, the European same-platform RGMB null is concordant between the targeted analysis and this unbiased scan. Eleventh, the cross-ancestry comparison was restricted to the 679 SomaScan v4 aptamers for which a significant cis-pQTL instrument was available in the Nagahama resource. Both resources use the SomaScan v4 platform, although the assayed aptamer sets are not identical (Nagahama: 4,392 aptamers analyzed, corresponding to 4,239 unique proteins; deCODE: 4,907 aptamers); the smaller number of usable instruments in Nagahama reflects its much smaller sample size (n ≈ 1,823 versus n ≈ 35,559 for deCODE) and the consequently lower power to detect cis-pQTLs, rather than a difference in the proteins measured. Restricting to these 679 shared instruments ensured a symmetric, like-for-like comparison, but means the East Asian– and European–enriched sets reflect only this shared panel. Proteins assayed in deCODE but lacking a Nagahama instrument (the deCODE resource yields significant cis-pQTLs for a far larger fraction of the panel) were not examined and could harbour additional European colocalizations, so the European–enriched set should not be read as a complete European proteome-wide profile. Finally, the Nagahama pQTL resource is based on an aptamer platform and, at the time of analysis, preprint-stage data; effect estimates and orientations should be reconfirmed against finalized releases. Three further limitations arise from the analyses added in revision. The lead variant was prioritized on the basis of its East Asian colocalization, so the East Asian effect estimate entering the cross-ancestry heterogeneity test may be inflated by selection, and that test quantifies the observed difference rather than confirming it independently. The multi-signal colocalization required removal of variants inconsistent with the reference LD and could not use coloc.susie, because SuSiE returned no credible set on the POAG side in either ancestry; conditioning and masking were used instead, and the conditional posterior in East Asians (PP4 = 0.35) was lower than the unconditional one (0.69), reflecting the weakness of the outcome signal. Finally, the trabecular meshwork data are from cultured cells rather than native tissue, are transcriptomic, and address the response to a single stimulus (TGF-β2), so the absence of a TGF-β2 response does not exclude regulation of RGMB by other stimuli relevant to outflow.

## Future Directions

Near-term, data-driven priorities follow directly from these limitations. Independent replication of the RGMB–POAG colocalization in additional East Asian POAG cohorts would test the robustness and the East Asian–enriched hypothesis. Acquisition of ancestry-matched ocular eQTL data, when available, would allow a tissue– and ancestry-matched expression colocalization that the present study could not perform. Orthogonal protein-level evidence (for example, curated retinal protein expression resources) could further contextualize the low retinal transcript abundance of RGMB.

Longer-term, functional validation in eye-relevant models would be required to move from a prioritized candidate toward a mechanistic target. Such work could include controlled exposure of human induced pluripotent stem cell (iPSC)-derived RGCs or retinal neuronal lines to recombinant RGMB with quantification of survival and neurite morphology; receptor-level perturbation (NEO1 or BMPR1A knockdown/knockout) to test pathway dependence; and tracking of downstream effectors (for example, SMAD1/5/8 and ROCK signaling) to characterize the mode of action. These experiments are proposed as hypotheses to be tested and are beyond the scope of the present data-driven analysis.

Beyond target nomination, the RGM–neogenin–BMP axis is pharmacologically tractable. Elezanumab, a clinical-stage human monoclonal antibody against the paralogous molecule RGMa, blocks RGM-mediated BMP/SMAD1/5/8 and neogenin signaling and, in optic nerve crush and optic neuritis models, preserves the retinal nerve fiber layer and promotes retinal ganglion cell survival^29,31^. Because higher plasma RGMB was associated with greater POAG risk in the present analysis, antagonism of RGMb is the direction-consistent hypothesis, although the dual role of RGMb as a BMP co-receptor and a PD-L2 receptor means that the net effect on retinal ganglion cells would require functional validation. RGMa and RGMb are paralogues that share roughly 50% amino-acid identity and a common neogenin-binding, BMP-co-receptor architecture, yet differ functionally (RGMb lacks the RGD motif, binds PD-L2, and has higher BMP2/4 affinity) and are not cross-targeted by a single antibody. Importantly, elezanumab targets RGMa rather than RGMb, no RGMb-selective agent has yet entered clinical development, and anti-RGMa efficacy remains clinically unproven, with negative primary endpoints in phase 2 multiple-sclerosis trials^32^; these observations therefore define a translational rationale rather than an established therapy.

## Conclusion

Integrating East Asian POAG and plasma pQTL data with multi-dataset and multi-signal colocalization, cross-ancestry fine-mapping, a formal test of effect heterogeneity, and expression mapping in retina and trabecular meshwork, and comparing colocalization profiles between Japanese and European populations on a common platform, we prioritize the RGMB–neogenin–BMP axis as a POAG candidate detected only in East Asian data. The signal does not colocalize with intraocular pressure in either ancestry, and the per-allele effect on POAG differs significantly between ancestries in data that are better powered on the European side. The findings are exploratory: they are consistent with an ancestry-enriched, pressure-independent locus but do not establish causality, ancestry specificity, or a link to the normal-tension phenotype, because the outcome GWAS was POAG rather than normal-tension glaucoma and the lead variant was prioritized in the same East Asian data. Ancestry-matched replication, tissue– and ancestry-matched expression data, and functional validation are the necessary next steps.

## Data Availability

All data analyzed in this study are publicly available summary-level datasets. POAG genetic associations were from BioBank Japan; the plasma cis-pQTL resource was from the Nagahama Study (SomaScan v4). LD reference data were from the 1000 Genomes Project Phase 3 (East Asian and European panels). The IGGC IOP GWAS is available via the GWAS Catalog (accession GCST009413); the Pan-UKBB IOP GWAS is available from the Pan-UKBB resource. The European POAG meta-analysis^15^ is available via the GWAS Catalog (accession GCST90011766), and the European plasma cis-pQTL data used for both the targeted RGMB analysis and the proteome-wide cross-ancestry scan are available from deCODE^14^. The IOP-independent POAG latent component^6^ is available from Zenodo. Single-cell retinal expression data were obtained from the CZ CELLxGENE Discover human retina atlas. Trabecular meshwork RNA-seq data were obtained from the Gene Expression Omnibus (accessions GSE236302 and GSE195756). Lead loci of the Japanese TMM IOP GWAS were used as published. Plasma proteomic (Olink Explore) associations from the China Kadoorie Biobank were accessed via the public CKB PheWeb portal (https://pheweb.ckbiobank.org). Analysis code is deposited at Zenodo (doi:10.5281/zenodo.23657164).

## Acknowledgments

The authors thank the participants and investigators of the studies and resources whose summary-level data made this work possible: BioBank Japan; the Nagahama Study; the Tohoku Medical Megabank Organization Eye Study; the China Kadoorie Biobank; deCODE genetics; the International Glaucoma Genetics Consortium; the Pan-UK Biobank team and the UK Biobank participants; the 1000 Genomes Project; the ENCODE Project Consortium; and the CZ CELLxGENE Discover platform. This work would not have been possible without their decision to make these resources publicly available.

Use of artificial intelligence: large-language-model–based tools (Claude; Anthropic, PBC, San Francisco, CA, USA; Claude Opus 4.8 and Claude Opus 5, accessed via the claude.ai interface between June and October 2026) were used to assist with analysis-code development, internal consistency checks, and manuscript drafting. No artificial-intelligence tool was used to evaluate or peer-review the work or to generate primary data. The authors reviewed and verified all analyses, text, and conclusions and take full responsibility for the content of the manuscript.

## Commercial relationships disclosures

The authors have no commercial relationships or conflicts of interest to disclose.

S. Okuno, None; S. Mizoue, None; K. Namiguchi, None; M. Yukawa, None; A. Shiraishi, None.

## Supplementary Material

**Supplementary Figure S1.** RGMB — cross-ancestry contrast in SMR, HEIDI and colocalization. (A) Colocalization posterior (PP4) of RGMB with POAG/IOP across the East Asian arm (Nagahama cis-pQTL × BBJ POAG) and European datasets. (B) Decomposition of the within-European colocalization posterior (deCODE RGMB pQTL × Gharahkhani POAG; 9,682 shared cis variants), which is dominated by H2 and H3 rather than H4. (C) Method-by-method summary for the two ancestry arms. SMR and HEIDI were applied in the East Asian arm only; the within-European analysis was colocalization-only by design.

**Supplementary Figure S2.** Concordance of the European arm with prior European proteome-wide MR/colocalization studies of POAG^7,8^. Paired colocalization posteriors (East Asian and European arms) for SVEP1, ITIH1, TMEM190 and RGMB, annotated with each protein’s status in the prior European studies; the recurrence of SVEP1 and ITIH1 supports the validity of the present scan.

**Supplementary Figure S3.** Protein-level concordance matrix across the present scan and prior European studies. Proteins are grouped into those recurring across studies (validity), present-study additions, prior-European targets assayed in the Nagahama panel but not colocalized (genuine cross-ancestry difference), and prior-European targets not represented in the 679-aptamer panel (coverage gap). Per-protein status in Yuan et al. (2024) and Jia et al. (2025) is shown alongside the present-study PP4 in each ancestry arm.

**Supplementary Figure S4.** Exploratory regulatory annotation of the RGMB rs1037599 LD block (chr5, GRCh38). East Asian LD proxies of rs1037599 (r² ≥ 0.8) are shown as ticks coloured by RegulomeDB rank. The lead variant rs1037599 (LD tag; rank 6) overlaps no ENCODE cCRE, whereas its proxy rs1563318 (r² = 0.84; rank 1b) lies within a distal enhancer-like cCRE (EH38E2394011); the dashed arc denotes a putative, unverified enhancer–promoter link. No single-TF motif disruption was detected (SNP2TFBS, 0/8).

**Supplementary Figure S5.** Regional plasma-RGMB cis-pQTL association at the RGMB locus (chr5, GRCh38), showing the lead/tag variant rs1037599 (purple diamond) and the regulatory candidate variant rs1563318 (red star; r² = 0.84), coloured by East Asian LD with rs1037599. rs1563318 lies ∼15 kb upstream of the lead at comparable association strength and within the same high-LD cluster; the RGMB gene body lies ∼320 kb downstream.

**Supplementary Table S1.** Status of prior-European POAG target proteins in the Nagahama 679-aptamer cis-pQTL panel, with colocalization PP4 in each ancestry arm and the in-panel/not-in-panel classification (generated by check_group_c.R; source file group_c_panel_status.csv).

**Supplementary Table S2.** p12 sensitivity of the RGMB colocalization between the China Kadoorie Biobank cis-pQTL and the Biobank Japan POAG GWAS. Posterior probabilities (PP.H0–PP.H4) from coloc.abf over 976 shared variants at prior probabilities of shared causality p12 = 1 × 10⁻⁵ (default), 1 × 10⁻⁶, and 1 × 10⁻⁷.

**Supplementary Table S3.** Direction of the RGMB rs1037599 cis-pQTL across three ancestries and two proteomic platforms, with full statistics (effect allele, per-allele effect on plasma RGMB, standard error, allele frequency, and cis-pQTL P) for Nagahama (Japanese, SomaScan v4), the China Kadoorie Biobank (Chinese, Olink Explore), and deCODE (European, SomaScan v4), together with the outcome-side direction of the RGMB-raising allele in Biobank Japan POAG.

**Supplementary Table S4. Datasets and parameters supplied to coloc.**

**Supplementary Table S5. Fine-mapping of the RGMB cis-region and linkage-disequilibrium diagnostics, by ancestry arm, cis-window and ridge regularization of the LD matrix.**

**Supplementary Table S6. Multi-signal colocalization of plasma RGMB with POAG, primary analysis (±500 kb, λ = 0).**

**Supplementary Table S7. Cross-ancestry heterogeneity in the per-allele POAG effect at rs1037599 and its 22 East Asian LD proxies (r² ≥ 0.8 in the 1000 Genomes East Asian panel).**

**Supplementary Table S8. RGMB-pathway, trabecular meshwork marker and control gene expression in primary human trabecular meshwork cells.**

**Supplementary Table S9. Summary-data–based Mendelian randomization (SMR) and HEIDI results for the 34 plasma proteins with SMR P < 0.05 and HEIDI P > 0.05 (Nagahama cis-pQTL × Biobank Japan POAG; 650 proteins tested), with the East Asian colocalization posterior (PP4) and the number of variants in each HEIDI test and colocalization.**

