## Supplementary materials for "Plasma Proteogenomics Identifies RGMB as a Candidate for Primary Open-Angle Glaucoma Detected Only in East Asians"

#### Supplementary Figures

**Supplementary Figure S1. RGMB — cross-ancestry contrast in SMR, HEIDI and colocalization.** (A) Colocalization posterior probability of a shared causal variant (PP4) of RGMB with POAG/IOP across the East Asian arm (Nagahama cis-pQTL × BBJ POAG) and European datasets. (B) Decomposition of the within-European colocalization posterior (deCODE RGMB pQTL × Gharahkhani POAG; 9,682 shared *cis* variants), which is dominated by H2 (signal in one dataset only) and H3 (distinct causal variants) rather than H4 (shared). (C) Method-by-method summary for the two ancestry arms. SMR and HEIDI were applied in the East Asian (discovery) arm only; the within-European analysis was colocalization-only by design. All present-study findings are exploratory (no protein survived Bonferroni correction).

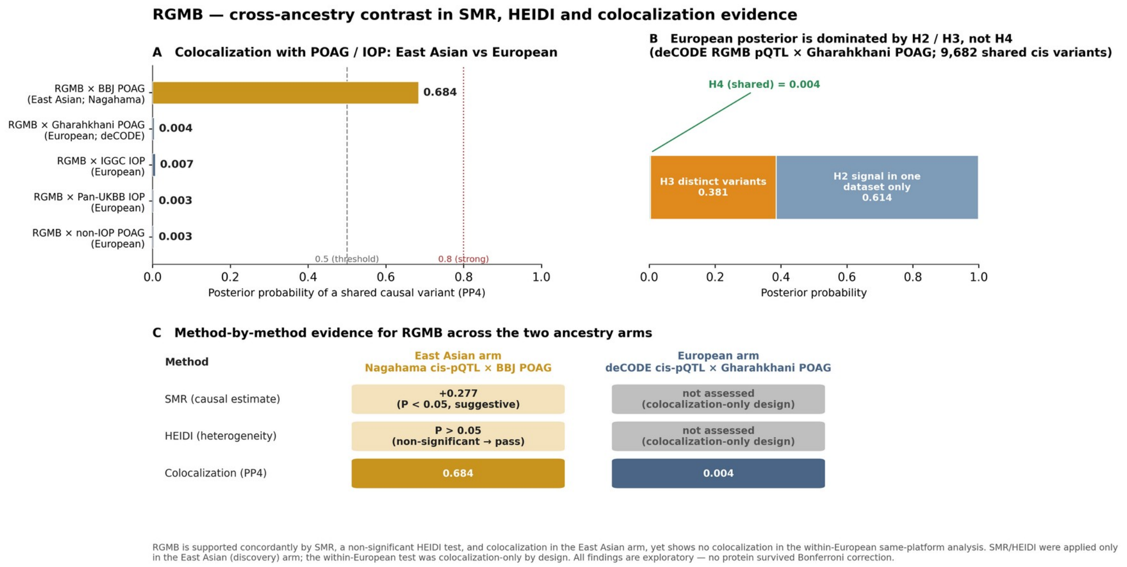


**Supplementary Figure S2. Concordance of the European arm with prior European proteome-wide MR/colocalization studies of POAG (Yuan et al., 2024; Jia et al., 2025).** Paired colocalization posteriors (East Asian and European arms) for SVEP1, ITIH1, TMEM190 and RGMB, annotated with each protein's status in the prior European studies. The recurrence of SVEP1 (a final European target in Jia et al., 2025) and ITIH1 (a European SMR-significant protein) supports the validity of the present scan; TMEM190 is outcome-

GWAS-dependent; RGMB is not a missed European signal (only the related RGMA appeared, weakly, and RGMA was not assayed in the Nagahama panel).

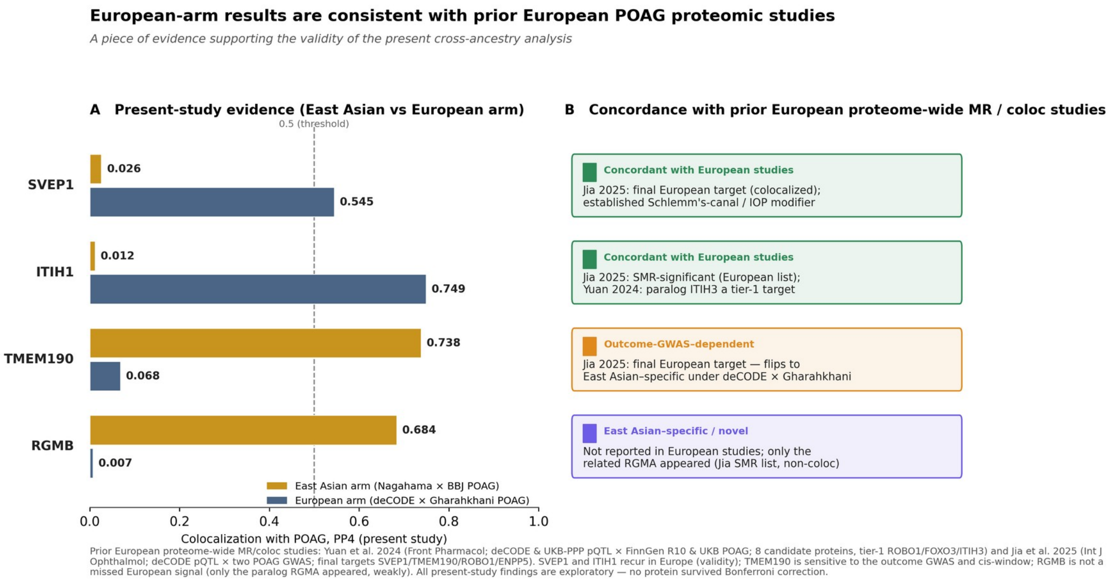


**Supplementary Figure S3. Protein-level concordance matrix across the present scan and prior European studies.** Proteins are grouped into: (A) recurring across studies (validity); (B) present-study additions not reported in European studies; (C1) prior-European targets assayed in the Nagahama 679-aptamer panel but not colocalized in either arm (a genuine cross-ancestry difference attributable to the outcome GWAS and method rather than to coverage); and (C2) prior-European targets not represented in the 679-aptamer panel (a coverage gap, not evaluable here). Per-protein status in Yuan et al. (2024; tier-1/tier-2 colocalization) and Jia et al. (2025; final target / SMR-significant) is shown alongside the present-study PP4 in each ancestry arm.

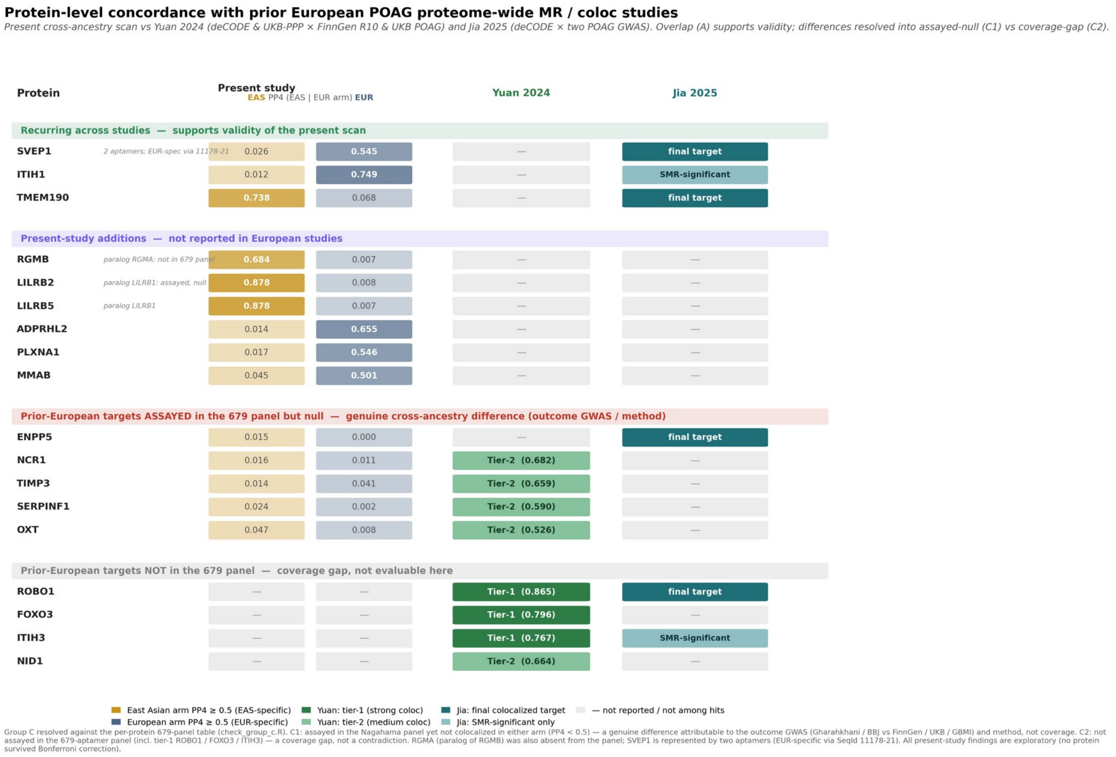


### Supplementary Tables

**Supplementary Table S1. Status of prior-European POAG target proteins in the Nagahama 679-aptamer cis-pQTL panel.** Colocalization PP4 is shown for each ancestry arm (East Asian: Nagahama cis-pQTL × BBJ POAG; European: deCODE cis-pQTL × Gharahkhani POAG). Classification uses PP4 ≥ 0.5. Generated by `check_group_c.R` (source file `group_c_panel_status.csv`).

| Protein | Prior-European source | In 679 panel | SeqId | PP4 (EAS) | PP4 (EUR) | Classification |
|---|---|---|---|---|---|---|
| ROBO1 | Yuan tier-1; Jia final | No | — | — | — | not in panel |
| FOXO3 | Yuan tier-1 | No | — | — | — | not in panel |
| ITIH3 | Yuan tier-1; Jia SMR-sig | No | — | — | — | not in panel |
| NID1 | Yuan tier-2 | No | — | — | — | not in panel |
| ENPP5 | Jia final | Yes | 6556-5 | 0.015 | 0.000 | neither (assayed, sub-threshold) |
| NCR1 | Yuan tier-2 | Yes | 8360- | 0.016 | 0.011 | neither (assayed, |

| Protein | Prior-European source | In 679 panel | SeqId | PP4 (EAS) | PP4 (EUR) | Classification |
|---|---|---|---|---|---|---|
| | | | 169 | | | sub-threshold) |
| TIMP3 | Yuan tier-2 | Yes | 2480-58 | 0.014 | 0.041 | neither (assayed, sub-threshold) |
| SERPINF1 | Yuan tier-2 | Yes | 9211-19 | 0.024 | 0.002 | neither (assayed, sub-threshold) |
| OXT | Yuan tier-2 | Yes | 8356-88 | 0.047 | 0.008 | neither (assayed, sub-threshold) |

*Sanity / paralog checks:* RGMA (paralog of RGMB) was not assayed in the panel; LILRB1 (paralog of LILRB2/LILRB5) was assayed but sub-threshold (SeqId 5090-49; PP4_EAS 0.044, PP4_EUR 0.021). SVEP1 is represented by two aptamers (SeqId 11178-21, EUR-enriched, PP4_EUR 0.545; SeqId 11109-56, sub-threshold). Ancestry-specific anchors reproduced as expected: ITIH1 (EUR-enriched, 0.749), TMEM190 (EAS-enriched, 0.738), RGMB (EAS-enriched, 0.684).

**Supplementary Table S2.** p12 sensitivity of the RGMB colocalization between the China Kadoorie Biobank cis-pQTL and the Biobank Japan POAG GWAS (coloc.abf, 976 shared variants). PP.H0 and PP.H2 were <0.001 at all priors and are shown as such.

| p12 | PP.H0 | PP.H1 | PP.H2 | PP.H3 | PP.H4 |
|---|---|---|---|---|---|
| $1 \times 10^{-5}$ (default) | <0.001 | 0.206 | <0.001 | 0.060 | 0.734 |
| $1 \times 10^{-6}$ | <0.001 | 0.607 | <0.001 | 0.177 | 0.216 |
| $1 \times 10^{-7}$ | <0.001 | 0.753 | <0.001 | 0.220 | 0.027 |

**Supplementary Table S3.** Direction of the RGMB rs1037599 cis-pQTL across three ancestries and two proteomic platforms. β is the per-G-allele effect on plasma RGMB on platform-specific scales (SomaScan, relative fluorescence units; Olink, normalized protein expression) and is not comparable in magnitude across platforms; only the effect allele and the sign are compared. The G allele raises plasma RGMB in all three cohorts.

| Cohort (ancestry) | Platform | Effect allele | β | SE | Allele freq | cis-pQTL P |
|---|---|---|---|---|---|---|
| Nagahama (Japanese) | SomaScan v4 | G | +0.300 | 0.036 | 0.35 | $9.8 \times 10^{-17}$ |
| CKB (Chinese) | Olink Explore | G | +0.180 | 0.024 | 0.39 | $2.9 \times 10^{-14}$ |
| deCODE (European) | SomaScan v4 | G | +0.124 | 0.008 | 0.47 | $2 \times 10^{-51}$ |

*Outcome direction (Biobank Japan POAG):* the RGMB-raising G allele is associated with higher POAG risk (log-odds ratio = +0.083; SE 0.026; $P = 1.5 \times 10^{-3}$).

**Supplementary Table S4.** Datasets and parameters supplied to coloc. Trait type, sample size and, for case–control outcomes, the case fraction were specified explicitly for every colocalization reported in this study. Approximate Bayes factors were computed from the per-variant effect estimate and its standard error. The variance of each quantitative exposure was set to unity because plasma protein levels had been rank-normalized in the source resources. Every colocalization was repeated at prior probabilities of shared causality p12 = $1 \times 10^{-5}$ (the coloc default), $1 \times 10^{-6}$ and $1 \times 10^{-7}$.

| Dataset | Role | Trait type | N | Case fraction | Exposure variance |
|---|---|---|---|---|---|
| Nagahama cis-pQTL (SomaScan v4) | Exposure | quantitative | 1,823 | — | 1 (rank-normalized) |
| deCODE cis-pQTL (SomaScan v4) | Exposure | quantitative | 35,559 | — | 1 (rank-normalized) |
| China Kadoorie Biobank (Olink) | Exposure | quantitative | ≈3,974 | — | 1 (rank-normalized) |
| BioBank Japan POAG | Outcome | case–control | 22,795 | 0.175 (3,980 cases) | — |
| Gharahkhani European POAG | Outcome | case–control | 216,257 | 0.0771 (16,677 cases) | — |
| IGGC IOP (GCST009413) | Outcome | quantitative | 31,269 | — | — |
| Pan-UKBB IOPcc | Outcome | quantitative | 87,199 | — | — |
| Non-IOP POAG latent component | Outcome | quantitative | nominal effective N | — | — |

**Supplementary Table S5.** Fine-mapping of the RGMB cis-region and linkage-disequilibrium diagnostics, by ancestry arm, cis-window and ridge regularization of the LD matrix. Variants inconsistent with the reference LD were removed before fine-mapping using the kriging_rss diagnostic of susieR (22 of 924 variants in the East Asian arm and 13 of 937 in the European arm at ±500 kb). The consistency estimate is estimate_s_rss; ρ is the Spearman correlation between $r^2$ with the strongest variant and |z|, which is expected to be positive when summary statistics and reference LD agree. λ = 0 is the primary analysis.

| Arm | cis-window | λ | Variants | estimate_s_rss | $\rho(r^2, \|z\|)$ | 95% credible sets | Top variant of leading set | PIP | Purity |
|---|---|---|---|---|---|---|---|---|---|

| | | | | | | | | | |
|---|---|---|---|---|---|---|---|---|---|
| East Asian | ±500 kb | 0 | 902 | 0.104 | 0.737 | 1 | rs1037599 | 0.269 | 0.880 |
| East Asian | ±250 kb | 0 | 483 | 0.116 | 0.705 | 1 | rs1037599 | 0.522 | 0.880 |
| East Asian | ±150 kb | 0 | 303 | 0.085 | 0.671 | 1 | rs1037599 | 0.589 | 0.911 |
| East Asian | ±500 kb | 0.01 | 902 | 0.095 | 0.737 | 1 | rs1037599 | 0.270 | 0.871 |
| East Asian | ±500 kb | 0.05 | 902 | 0.057 | 0.737 | 1 | rs1037599 | 0.272 | 0.836 |
| European | ±500 kb | 0 | 924 | 0.086 | 0.695 | 10 | chr5:98,730,764 | 1.000 | 1.000 |
| European | ±250 kb | 0 | 497 | 0.073 | 0.654 | 7 | chr5:98,506,285 | 1.000 | 1.000 |
| European | ±150 kb | 0 | 316 | 0.050 | 0.554 | 7 | chr5:98,506,285 | 1.000 | 1.000 |
| European | ±500 kb | 0.01 | 924 | 0.077 | 0.695 | 10 | chr5:98,730,764 | 1.000 | 1.000 |
| European | ±500 kb | 0.05 | 924 | 0.038 | 0.695 | 10 | chr5:98,730,764 | 1.000 | 1.000 |

Before outlier removal the consistency estimate was 0.80 in the East Asian arm and 0.62 in the European arm, and SuSiE did not converge to a stable prior variance in either. All fine-mapping reported here converged with the default prior check enabled. SuSiE applied to the POAG side returned no 95% credible set in either ancestry, reflecting the limited power of the outcome GWAS at this locus; coloc.susie therefore could not be used, and conditioning and masking were used in its place.

**Supplementary Table S6.** Multi-signal colocalization of plasma RGMB with POAG, primary analysis (±500 kb, λ = 0). coloc.abf assumes at most one causal variant per trait; coloc.signals relaxes that assumption by masking variants in linkage disequilibrium with a resolved signal, or by conditioning on it. Where a method resolved more than one signal pair, the largest PP4 across pairs is shown. Full per-pair posteriors at all three priors are deposited at Zenodo.

| Arm | Method | Signal pairs | PP4 (p12 = $1 \times 10^{-5}$) | PP4 ($1 \times 10^{-6}$) | PP4 ($1 \times 10^{-7}$) | Dominant alternative |
|---|---|---|---|---|---|---|
| East Asian | coloc.abf | 1 | 0.692 | 0.183 | 0.022 | PP.H1 = 0.255 |
| East Asian | coloc.signals, masking | 1 | 0.691 | 0.183 | 0.022 | PP.H1 = 0.254 |
| East Asian | coloc.signals, conditioning | 1 | 0.350 | 0.051 | 0.005 | PP.H1 = 0.622 |
| European | coloc.abf | 1 | 0.0068 | 0.00069 | 0.000069 | PP.H1 = 0.964 |
| European | coloc.signals, masking | 3 | 0.017 | 0.0017 | 0.00017 | PP.H1 = 0.955–0.970 |
| European | coloc.signals, | 6 | 0.000024 | 0.0000024 | 0.00000024 | PP.H3 = 0.952– |

| conditioning | 1.000 |
|---|---|

In the East Asian arm every method resolved a single signal pair led by rs1037599, and the alternative favoured by the conditional analysis was PP.H1 — a signal in the pQTL data only — rather than PP.H3, so the lower conditional posterior reflects the weakness of the outcome signal rather than evidence for distinct causal variants (PP.H3 = 0.027). In the European arm conditioning favoured PP.H3, that is, it positively supported distinct causal variants rather than merely failing to support a shared one. Across the sensitivity analyses (±250 kb, ±150 kb, λ = 0.01) the European posteriors remained below 0.02 except for a single masked signal pair at ±150 kb (PP4 = 0.19).

**Supplementary Table S7.** Cross-ancestry heterogeneity in the per-allele POAG effect at rs1037599 and its 22 East Asian LD proxies ($r^2$ ≥ 0.8 in the 1000 Genomes East Asian panel). Effects are aligned to the plasma-RGMB-raising allele. Q is Cochran's Q with one degree of freedom; P values were adjusted across the 22 variants by the Benjamini–Hochberg procedure (all FDR < 0.01). † lead variant; ‡ the regulatory candidate within a distal enhancer-like cCRE. Positions (GRCh38) and the full output are deposited at Zenodo.

| Variant | Effect allele | β (SE), East Asian | P | β (SE), European | P | Q | P (Q) | $I^2$ (%) |
|---|---|---|---|---|---|---|---|---|
| rs1037599 † | G | +0.083 (0.026) | 0.0015 | −0.0016 (0.0129) | 0.904 | 8.50 | 0.0036 | 88.2 |
| rs6870628 | T | +0.090 (0.026) | 0.00053 | +0.0054 (0.0129) | 0.678 | 8.50 | 0.0036 | 88.2 |
| rs11241450 | G | +0.083 (0.026) | 0.0015 | −0.0014 (0.0129) | 0.915 | 8.46 | 0.0036 | 88.2 |
| rs1037598 | C | +0.083 (0.026) | 0.0015 | −0.0010 (0.0129) | 0.936 | 8.38 | 0.0038 | 88.1 |
| rs12152757 | T | +0.083 (0.026) | 0.0015 | −0.0010 (0.0129) | 0.937 | 8.38 | 0.0038 | 88.1 |
| rs9686816 | C | −0.088 (0.026) | 0.00070 | −0.0045 (0.0129) | 0.727 | 8.28 | 0.0040 | 87.9 |
| rs1037597 | C | +0.087 (0.026) | 0.00077 | +0.0043 (0.0129) | 0.738 | 8.12 | 0.0044 | 87.7 |
| rs10035465 | T | +0.087 (0.026) | 0.00076 | +0.0043 (0.0129) | 0.741 | 8.12 | 0.0044 | 87.7 |
| rs1072722 | A | −0.087 (0.026) | 0.00076 | −0.0046 (0.0129) | 0.720 | 8.06 | 0.0045 | 87.6 |
| rs7717913 | C | +0.087 (0.026) | 0.00076 | +0.0046 (0.0129) | 0.722 | 8.06 | 0.0045 | 87.6 |
| rs7717448 | C | +0.087 (0.026) | 0.00076 | +0.0047 (0.0129) | 0.714 | 8.04 | 0.0046 | 87.6 |
| rs10063500 | C | +0.087 (0.026) | 0.00077 | +0.0048 (0.013) | 0.711 | 8.00 | 0.0047 | 87.5 |
| rs751882 | C | +0.087 (0.026) | 0.00077 | +0.0049 (0.013) | 0.708 | 7.98 | 0.0047 | 87.5 |
| rs723207 | T | +0.087 | 0.00076 | +0.0051 | 0.696 | 7.96 | 0.0048 | 87.4 |

| | | | | | | | | |
|---|---|---|---|---|---|---|---|---|
| | | (0.026) | | (0.0129) | | | | |
| rs723206 | T | +0.087 (0.026) | 0.00076 | +0.0053 (0.0129) | 0.682 | 7.92 | 0.0049 | 87.4 |
| rs6865661 | G | −0.087 (0.026) | 0.00077 | −0.0052 (0.013) | 0.688 | 7.92 | 0.0049 | 87.4 |
| rs1002268 | C | +0.088 (0.026) | 0.00077 | +0.0072 (0.013) | 0.579 | 7.73 | 0.0054 | 87.1 |
| rs1563317 | G | +0.086 (0.026) | 0.00089 | +0.0053 (0.013) | 0.685 | 7.71 | 0.0055 | 87.0 |
| rs59060802 | C | +0.086 (0.026) | 0.00083 | +0.0061 (0.013) | 0.637 | 7.56 | 0.0060 | 86.8 |
| rs1563318 ‡ | G | +0.086 (0.026) | 0.00083 | +0.0061 (0.013) | 0.639 | 7.56 | 0.0060 | 86.8 |
| rs3890433 | T | −0.086 (0.026) | 0.00083 | −0.0064 (0.013) | 0.620 | 7.50 | 0.0062 | 86.7 |
| rs11241481 | T | +0.083 (0.026) | 0.0016 | +0.0073 (0.0129) | 0.571 | 6.80 | 0.0091 | 85.3 |

The asymmetry in sample size favours the European dataset on both sides of the analysis: the European POAG meta-analysis has 4.2 times as many cases and a standard error half as large, and the European cis-pQTL instrument is the more precisely estimated of the two ($z = 15.1$ versus 8.4). The European 95% confidence interval at the lead variant (−0.027 to +0.024) excludes the East Asian point estimate, and an effect of the East Asian magnitude would have been detected in the European data at $Z = 6.4$ ($P = 1.2 \times 10^{-10}$). Because rs1037599 was prioritized on the basis of its colocalization in the East Asian data, the East Asian effect estimate may be subject to selection bias, and this test should be read as a quantification of the observed difference rather than as independent confirmation.

**Supplementary Table S8.** RGMB-pathway, trabecular meshwork marker and control gene expression in primary human trabecular meshwork cells. GSE236302 comprises eight donors (untreated control versus TGF-β2; seven donors contributed both conditions); log2 fold changes are from paired t tests on log2 CPM with Benjamini–Hochberg correction across the panel. The percentile is the rank among the 15,973 genes with mean CPM > 1, so that abundance is judged against the expressed transcriptome rather than against all annotated genes. The final column gives the per-sample CPM range in the independent dataset GSE195756 (eight samples), which was used to confirm expression only.

| Gene | Mean CPM | Percentile among expressed genes | log2 FC (TGF-β2 vs control) | P | FDR | CPM range, GSE195756 |
|---|---|---|---|---|---|---|
| RGMB | 22.1 | 52 | −0.001 | 0.997 | 0.997 | 25.4–95.7 |
| NEO1 | 27.5 | 57 | −0.036 | 0.805 | 0.898 | 55.8–211.7 |
| BMPR1A | 70.9 | 79 | −0.037 | 0.684 | 0.826 | 28.7–81.6 |
| BMPR2 | 85.2 | 83 | +0.477 | 0.012 | 0.026 | 78.3–204.1 |
| BMPR1B | 11.4 | 40 | +0.572 | 0.0018 | 0.0066 | — |
| BMP2 | 238 | 96 | +1.50 | $3.9 \times 10^{-9}$ | $1.1 \times 10^{-7}$ | — |

| | | | | | | |
|---|---|---|---|---|---|---|
| BMP4 | 32.0 | 61 | −1.45 | 0.0018 | 0.0066 | — |
| RGMA | 2.79 | 18 | −0.478 | 0.013 | 0.026 | — |
| MYOC (TM marker) | 472 | 98.5 | −1.19 | 0.042 | 0.068 | — |
| CHI3L1 (TM marker) | 803 | 99.3 | −2.37 | 0.0012 | 0.0066 | — |
| TGFB2 | 15.0 | 44 | +0.194 | 0.331 | 0.439 | — |
| TGFBR1 | 120 | 88 | +0.809 | 0.0041 | 0.013 | — |
| TGFBR2 | 222 | 95 | −0.810 | 0.00082 | 0.0066 | — |
| SERPINE1 (TGF-β2 target) | 701 | 99.1 | +1.89 | $2.5 \times 10^{-4}$ | 0.0037 | — |
| ACTA2 (TGF-β2 target) | 404 | 98.1 | +1.24 | 0.0063 | 0.018 | — |
| COL1A1 (TGF-β2 target) | 177 | 93 | +1.12 | 0.013 | 0.026 | — |
| FN1 (TGF-β2 target) | 919 | 99.4 | +0.861 | 0.057 | 0.083 | — |
| RHO (negative control) | 0.04 | not expressed | −0.012 | 0.594 | 0.748 | — |

TGF-β2 treatment produced the expected fibrotic response, yet RGMB, NEO1 and BMPR1A were unchanged. BMP ligands did shift (BMP2 +1.50, BMP4 −1.45), consistent with the reported antagonism between BMP-4 and TGF-β2 in the trabecular meshwork. RBPMS, used as a retinal ganglion cell marker elsewhere in this study, is expressed broadly outside the retina and is therefore not a negative control in this tissue; RHO serves that role and was absent (detected in 33% of samples at 0.04 CPM). These data derive from cultured cells rather than native tissue and are transcriptomic, so they address the presence of the pathway and its response to a single stimulus rather than its activity in vivo.

**Supplementary Table S9.** Summary-data–based Mendelian randomization (SMR) and HEIDI results for the 34 plasma proteins with SMR P < 0.05 and HEIDI P > 0.05 (Nagahama cis-pQTL × Biobank Japan POAG; 650 proteins tested), ordered by SMR P. b_SMR is the estimated effect on POAG (log-odds) per unit increase in standardized plasma protein level; HEIDI variants is the number of variants used in the HEIDI test. PP4 is the posterior probability of a shared causal variant from coloc.abf (default priors) over the cis-region. Proteins with PP4 ≥ 0.5 are shown in bold, with their gene names.

| SomaScan SeqId | Top cis-pQTL (GRCh38) | b_SMR (SE) | SMR P | HEIDI P | HEIDI variants | PP4 (East Asian) | Variants in coloc |
|---|---|---|---|---|---|---|---|
| **10442-1 (TMEM190)** | **19:55376727:C:T** | **-0.086 (0.026)** | **1.1e-03** | **0.87** | **20** | **0.738** | **997** |
| **3331-8 (RGMB)** | **5:98447787:T:G** | **+0.277 (0.093)** | **2.9e-03** | **0.95** | **20** | **0.684** | **1010** |
| 11208-15 | 16:5030120:C:T | -0.144 (0.049) | 3.5e-03 | 0.23 | 20 | 0.464 | 1131 |
| 3352-80 | 1:8949392:G:C | +0.132 (0.048) | 6.0e-03 | 0.19 | 16 | 0.392 | 957 |
| 5708-1 | 5:132860691:A:C | +0.238 (0.087) | 6.3e-03 | 0.79 | 10 | 0.285 | 1014 |

| | | | | | | | |
|---|---|---|---|---|---|---|---|
| 6617-12 | 1:159812894:A:G | -0.236 (0.088) | 7.3e-03 | 0.32 | 10 | 0.422 | 1353 |
| 5491-12 | 10:72106753:T:C | -0.296 (0.112) | 8.5e-03 | 0.72 | 16 | 0.398 | 1378 |
| 19277-4 | 1:161042970:T:C | -0.206 (0.078) | 8.7e-03 | 0.99 | 20 | 0.311 | 1052 |
| 12387-7 | 5:132271243:G:A | -0.179 (0.068) | 8.8e-03 | 0.47 | 20 | 0.322 | 974 |
| 15686-49 | 12:57433199:A:G | -0.078 (0.030) | 0.01 | 0.73 | 20 | 0.322 | 691 |
| **7015-8 (LILRB5)** | **19:54250521:T:G** | **-0.105 (0.041)** | **0.01** | **0.48** | **5** | **0.878** | **690** |
| 18289-16 | 17:36001998:T:G | +0.055 (0.021) | 0.01 | 0.29 | 17 | 0.241 | 695 |
| 9370-69 | 8:63027655:C:T | +0.082 (0.033) | 0.01 | 0.15 | 20 | 0.190 | 1390 |
| 8024-64 | 22:26520756:C:T | -0.262 (0.108) | 0.01 | 0.79 | 17 | 0.259 | 1754 |
| 9313-27 | 16:48965278:A:T | -0.060 (0.025) | 0.02 | 0.52 | 12 | 0.175 | 1286 |
| 4541-49 | 11:126019569:A:G | +0.102 (0.044) | 0.02 | 0.18 | 18 | 0.186 | 1455 |
| 4459-68 | 11:117211047:C:G | -0.056 (0.024) | 0.02 | 0.22 | 20 | 0.141 | 1700 |
| 13624-17 | 1:1844830:C:A | +0.233 (0.104) | 0.02 | 0.91 | 15 | 0.159 | 733 |
| 4435-66 | 17:79724812:T:C | -0.059 (0.026) | 0.03 | 0.51 | 9 | 0.119 | 1498 |
| 7227-75 | 14:30782789:G:A | +0.197 (0.089) | 0.03 | 0.71 | 6 | 0.212 | 1041 |
| 10714-7 | 17:63493061:A:G | -0.078 (0.036) | 0.03 | 0.72 | 12 | 0.113 | 549 |
| 8007-19 | 8:11844497:A:T | +0.176 (0.080) | 0.03 | 0.98 | 20 | 0.217 | 1018 |
| 9950-229 | 12:6775910:G:A | +0.203 (0.094) | 0.03 | 0.82 | 8 | 0.104 | 1046 |
| 3431-54 | 7:143406287:T:C | -0.217 (0.101) | 0.03 | 0.77 | 11 | 0.124 | 465 |
| 18864-7 | 9:33958346:A:G | +0.121 (0.057) | 0.03 | 0.20 | 20 | 0.094 | 596 |
| 13405-61 | 4:56824645:C:G | -0.114 (0.054) | 0.04 | 0.55 | 20 | 0.145 | 1541 |
| 8606-39 | 7:23273552:T:C | +0.083 (0.039) | 0.04 | 0.61 | 20 | 0.096 | 1057 |
| 12378-71 | 6:33323048:A:G | -0.120 (0.058) | 0.04 | 0.39 | 20 | 0.001 | 2198 |
| 4337-49 | 1:159724989:T:C | +0.238 (0.116) | 0.04 | 0.64 | 13 | 0.110 | 1363 |
| 6580-29 | 12:9164311:C:T | -0.119 (0.059) | 0.04 | 0.06 | 20 | 0.085 | 987 |
| 6425-87 | 12:55992292:A:G | -0.233 (0.116) | 0.05 | 0.38 | 5 | 0.189 | 726 |
| 10818-36 | 11:6394233:G:A | -0.142 (0.071) | 0.05 | 0.71 | 7 | 0.109 | 1503 |
| 4324-33 | 20:23746251:C:T | -0.126 (0.063) | 0.05 | 0.66 | 5 | 0.125 | 1602 |
| 15333-11 | 17:28344170:C:G | -0.115 (0.058) | 0.05 | 0.46 | 17 | 0.087 | 440 |

No protein survived Bonferroni correction across the 650 proteins tested. Of the 34, three also colocalized with POAG (PP4 ≥ 0.5): LILRB5, TMEM190 and RGMB. LILRB5 was not prioritized because its HEIDI test rested on five variants and its colocalization (PP4 = 0.878) was indistinguishable from that of the neighbouring LILRB2 (PP4 = 0.878), which was not among the 34.

**Supplementary Note: exploratory regulatory annotation of rs1037599.** These analyses are purely exploratory and hypothesis-generating, and were not used for candidate prioritization: all supporting chromatin and transcription-factor evidence derives from non-ocular ENCODE biosamples, no allele-specific functional assay was performed, and the physical enhancer–RGMB link was not tested. Because the RGMB lead pQTL variant rs1037599 lies ~320 kb upstream of the RGMB gene body and was not resolved as the causal variant in fine-mapping, we annotated rs1037599 and its 40 East Asian LD proxies ($r^2 \geq 0.8$; 1000 Genomes EAS) against ENCODE SCREEN candidate cis-regulatory elements (cCREs) and RegulomeDB, and screened the eight variants that fell within the two candidate cCRE sub-regions for allele-specific transcription-factor (TF) binding-site matches with SNP2TFBS. rs1037599 itself did not overlap any cCRE and received a low regulatory score (RegulomeDB rank 6), arguing against a direct regulatory role. In contrast, an LD proxy, rs1563318 ($r^2 = 0.84$ with rs1037599), fell within a distal enhancer-like cCRE (EH38E2394011; H3K4me3 + H3K27ac, DNase max-Z = 4.1) and carried strong regulatory evidence (RegulomeDB rank 1b: TF ChIP, chromatin accessibility, and footprint support), with its plasma-RGMB-increasing allele (G) matching the pQTL direction. Regulatory evidence in this block clustered into two sub-regions (~98.432 Mb and ~98.454 Mb, GRCh38), the former coinciding with the nearest annotated cCREs. However, none of these eight variants overlapped a SNP2TFBS transcription-factor binding-site model (0 of 8 matched), so no allele-specific single-motif change could be scored; rs1037599 itself was not motif-tested, as it overlaps no cCRE. This lack of a scorable match is not positive evidence against a regulatory effect; it indicates only that the association is not explained by disruption of a single dominant motif represented in this model set. All supporting chromatin and TF evidence derives from non-ocular ENCODE biosamples, and the physical enhancer-RGMB link was not tested (no eye-tissue 3C/Hi-C). These exploratory analyses are hypothesis-generating: they indicate that the RGMB association most likely tags a regulatory module marked by rs1563318 rather than acting through rs1037599 itself, consistent with the cautious “distal upstream regulatory candidate, or a tag for one” interpretation in the main text. Functional and ancestry/tissue-matched data will be required to confirm the responsible element, variant, and transcription factor(s). These variant-level annotations are consistent with the SMR and colocalization results, which operate at the level of the RGMB cis-region rather than resolving individual variants: SMR used the lead cis-pQTL (rs1037599) as the instrument, and coloc.abf evaluated the entire cis window (lead variant ± 1 Mb) under a single-causal-variant assumption, so it cannot distinguish variants in high linkage disequilibrium. rs1563318 lies within the same ~27 kb high-LD block ($r^2 = 0.84$ with rs1037599) captured by the colocalization signal (PP4 = 0.684); the regulatory annotation therefore refines, rather than contradicts, the colocalization evidence by nominating a regulatory candidate within that block.

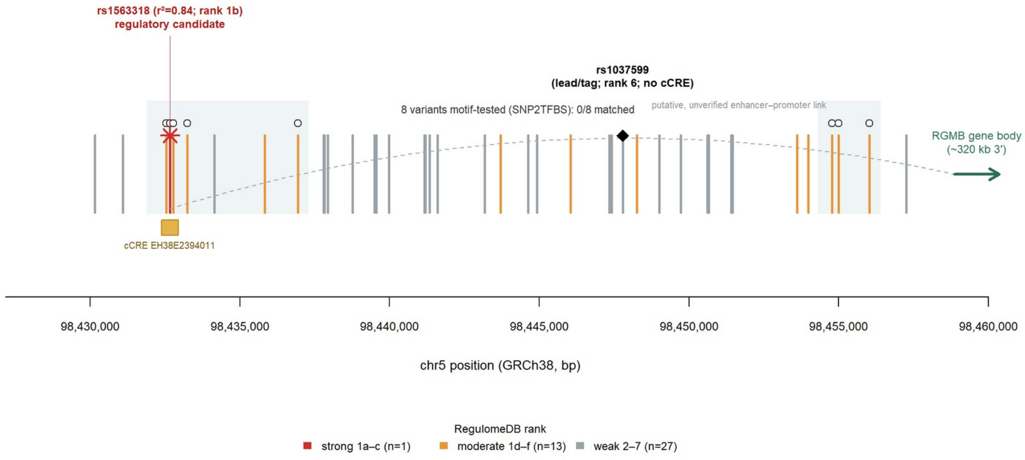


**Supplementary Figure S4.** Exploratory regulatory annotation of the RGMB rs1037599 LD block (chr5, GRCh38). The 40 variants in East Asian LD with rs1037599 ($r^2 \geq 0.8$; 41 including the lead) are shown as ticks coloured by RegulomeDB rank in three tiers: strong (1a–c; n = 1; rs1563318, rank 1b), moderate (1d–f; n = 13), and weak (2–7; n = 27). The lead pQTL variant rs1037599 (LD tag; rank 6) does not overlap any ENCODE cCRE, whereas its proxy rs1563318 ($r^2 = 0.84$; rank 1b; star) lies within a distal enhancer-like cCRE (EH38E2394011; gold box). The RGMB gene body lies ~320 kb downstream (right; axis break); the dashed arc denotes a purely schematic and unverified enhancer–promoter link. Of these, the eight variants within the two candidate cCRE sub-regions were queried in SNP2TFBS; none overlapped a modelled transcription-factor binding site (0/8 matched), so no single-motif change could be scored.

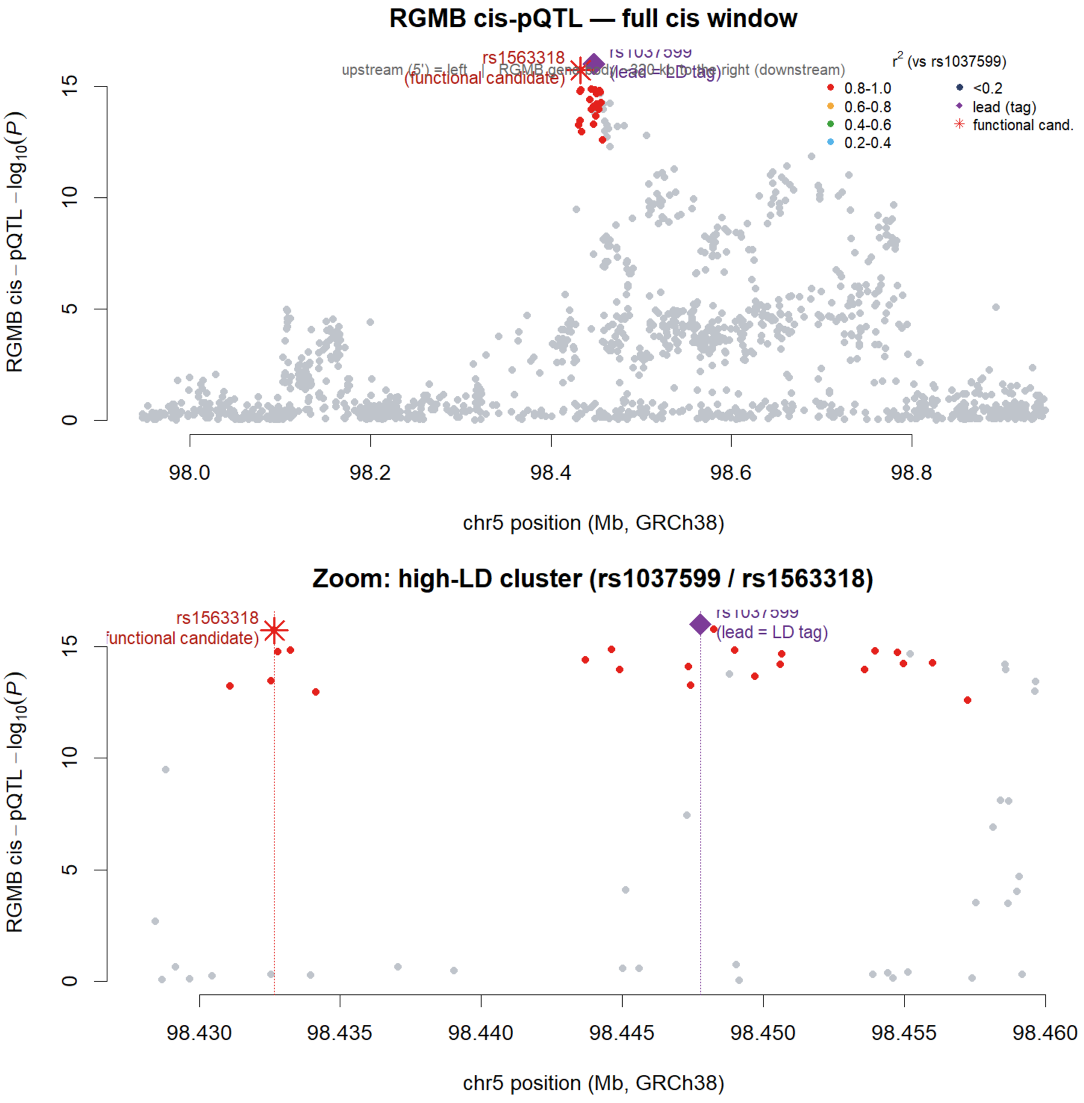


**Supplementary Figure S5.** Regional plasma-RGMB cis-pQTL association at the RGMB locus, showing the lead/tag and regulatory candidate variants (chr5, GRCh38). Each point is a cis variant plotted by genomic position (x-axis) against its association with plasma RGMB ($-\log_{10}P$, y-axis), coloured by East Asian LD ($r^2$) with the lead variant rs1037599. (Top) Full cis window; (bottom) zoom on the high-LD cluster. The lead pQTL variant rs1037599 (purple diamond) is the LD tag, and its proxy rs1563318 (red star; $r^2 = 0.84$) — the regulatory candidate variant (Supplementary Note; Supplementary Figure S4) — lies ~15 kb upstream (5′) of the lead at a comparable association strength ($-\log_{10}P \approx 15.7$ versus 16.0) and within the same high-LD cluster (red points). The RGMB gene body lies ~320 kb to the right (downstream). The figure visualises, on the pQTL association landscape, that the regulatory candidate rs1563318 sits among the top variants alongside the lead rather than as an isolated signal.
